# Structural evolution of trypsinogen gene redundancy confers risk for pancreas diseases

**DOI:** 10.1101/2022.08.08.22278454

**Authors:** Haiyi Lou, Yimin Wang, Bo Xie, Xinyue Bai, Yang Gao, Rui Zhang, Shuhua Xu

**Affiliations:** State Key Laboratory of Genetic Engineering, Center for Evolutionary Biology, Collaborative Innovation Center of Genetics and Development, School of Life Sciences, Fudan University, Shanghai 200438, China; Key Laboratory of Computational Biology, Shanghai Institute of Nutrition and Health, University of Chinese Academy of Sciences, Chinese Academy of Sciences, Shanghai 200031, China; School of Life Science and Technology, ShanghaiTech University, Shanghai 201210, China; Human Phenome Institute, Zhangjiang Fudan International Innovation Center, and Ministry of Education Key Laboratory of Contemporary Anthropology, Fudan University, Shanghai 201203, China; Department of Liver Surgery and Transplantation Liver Cancer Institute, Zhongshan Hospital, Fudan University, Shanghai 200032, China; Center for Excellence in Animal Evolution and Genetics, Chinese Academy of Sciences, Kunming 650223, China

**Keywords:** Trypsinogen gene, gene duplication, structural variation, pseudogene, *PRSS1-PRSS2*

## Abstract

Trypsin is an important enzyme secreted by the pancreas for digesting proteins. The precursors of major human trypsin are encoded by trypsinogen genes *PRSS1* and *PRSS2*. Here, we leveraged multi-omic data to study their evolutionary and functional impact. We estimated that the primate trypsinogen gene was duplicated from a single copy to multiple-copy 24-34 million years ago (Mya). Compared to six protein-coding genes in non-human great apes, the human ancestral state was a 5-copy with three being pseudogenized. Interestingly, a derived 3-copy form emerged in Africans ∼260 Kya and dominated in non-Africans as one of the two major haplotypes. Although no longer encoding proteins, the pseudogene enhancers still function on pancreatic *PRSS2* expression, leading to ∼15% up-regulation for the 5-copy than the 3-copy haplotype. Notably, the 3-copy structure was under positive selection in East Asians, where lower trypsin might be adaptive during high-starch diet shift for protecting the pancreas from autodigestion, as also supported by the identified causality of the haplotype structure to pancreatitis risk. Our efforts in elucidating the structural evolution of trypsinogen genes advance our understanding of the genetic basis and molecular mechanism of human pancreas diseases.

## Introduction

Trypsin is a digestive enzyme with the function of cutting peptide chains at the carboxyl group of lysine or arginine. It is activated in the small intestine from its precursor form trypsinogen, an inactive protease secreted by the pancreas. In humans, there are three major trypsinogen isoforms, cationic trypsinogen, anionic trypsinogen, and mesotrypsinogen. The cationic and anionic trypsinogen contribute more than 90% of the human trypsinogen in pancreas juice ^1,2^, which are encoded by *PRSS1* and *PRSS2* at the trypsinogen gene cluster on chromosome 7, respectively.

The DNA sequence structure of the human *PRSS1-PRSS2* locus was first discovered as five homologous trypsinogen genes (5-copy structure) flanking by the β T cell receptor genes ^3^. Later, a common deletion (3-copy structure) at this locus was reported ^4^. Both of the two structural forms of trypsinogen genes were included in the human reference genome GRCh38, with the 3-copy structure in the primary scaffold and the 5-copy structure in the alternative contig (Fig. 1a). These trypsinogen genes are located tandemly and each copy has a length of ∼10.6-kb and a high similarity (∼90%) with other trypsinogen gene copies. The first and the last copy (the number following the order of centromeric to telomeric positions) in both forms are protein-coding genes *PRSS1* and *PRSS2*, respectively, while the rest copies are all pseudo-genes. Although early studies investigated the evolution of peptide sequences in multiple species ^5,6^, the evolutionary history and biological impact of the trypsinogen gene duplication remain unknown.

**Fig. 1.**
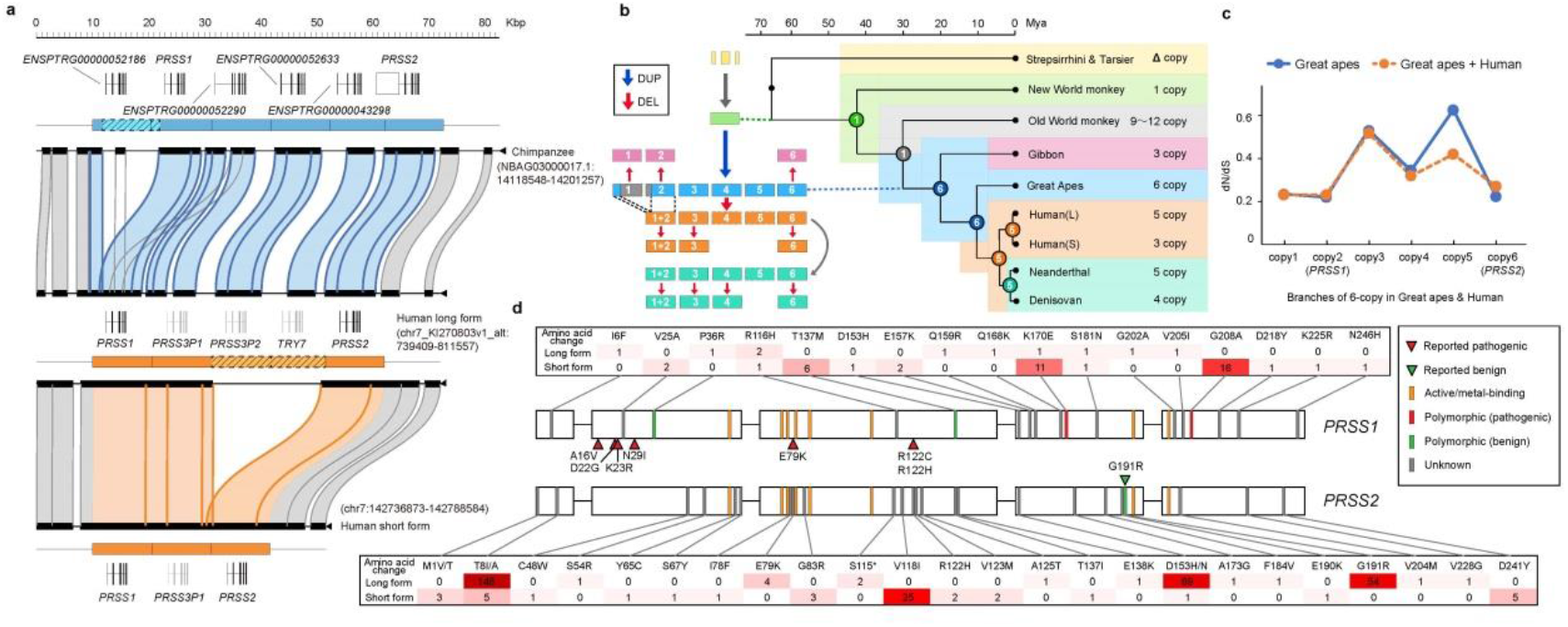
Trypsinogen gene evolution in primates. **a**. Trypsinogen gene sequence structure of tandem duplications at *PRSS1-PRSS2* locus in chimpanzee (6-copy, upper panel) and human (5-copy, lower panel). **b**. Phylogenetic tree with copy number of trypsinogen genes at *PRSS1-PRSS2* locus in primates. The circles at the internal nodes denote the inferred copy number of the common ancestry. The diagram denotes the evolutionary copy number change in ape lineage. **c**. Branch test of dN/dS ratio for different trypsinogen gene copies in nonhuman great apes (blue) and great apes (orange). See phylogeny of the branch test in Supplementary Fig. 7a and 7b. **d**. Amino acid changes of *PRSS1* and *PRSS2* found in 2,027 worldwide human samples. The shaded colors and the numbers in the box denote the count of haplotypes.

Due to the protein-digestive nature, excessive activation of trypsinogen might lead to autodigestion, an injury to the gland, which is one of the mechanisms for pancreatitis ^7^. Mutations in the trypsinogen genes might lead to pancreas diseases. Previous studies have found that the missense variants on *PRSS1* and triplication of the entire *PRSS1-PRSS2* locus would cause pancreatitis ^8,9^, but none of the pathogenic amino acid changes on *PRSS2* was reported ^10,11^, which implies that natural selection constraint varies among different trypsinogen genes. In addition, a genome-wide association study (GWAS) reported a common variant (rs10273639) associated with the risk of chronic pancreatitis at the *PRSS1-PRSS2* locus ^12^. Further efforts were made to search for causal variants linked to rs10273639, such as a variant reported being associated with *PRSS1* promotor activity ^13^. However, this variant does not account for the common variants to affect *PRSS1* expression in the pancreas tissue ^14^, which leaves the causality of the common variants contributing to pancreatitis predisposition remains elusive.

To address these problems, we reconstructed the evolutionary history of trypsinogen genes in primates and analyzed the haplotype polymorphism in humans for more than 2,000 worldwide samples. Leveraging multi-omics data, we explored the functional impact of the haplotype structure and its association with pancreas disease. Our analyses provided an evolutionary view of how human trypsinogen genes evolved and their implication in pancreas disease.

## Results

### Trypsinogen gene duplication in the primates

To gain an evolutionary insight into trypsinogen gene duplications at the *PRSS1-PRSS2* locus, we investigated the homologous sequences in primates (Supplementary Fig. 1 and 2). The primate trypsinogen genes started with one copy as shown in partial alignment with Strepsirrhini, and complete alignment of an entire copy was observed in the new world monkeys (Fig. 1b; Supplementary Fig. 3a; Supplementary Note). The gene duplications were found in both the old world monkey (9-12 copies) and the ape (3-6 copies) lineages. The Bayesian phylogenetic analysis showed that the duplication events might occur independently after the divergence of these two lineages, with an early rapid expansion during 31-34 Mya in the old-world monkeys and a later one during 24-29 Mya in the apes (Supplementary Fig. 3b), both within Oligocene Epoch of the Paleogene Period. In each of the two lineages, the duplication phylogeny follows a pattern that the orthologous genes of different species cluster together rather than the paralogous genes of the same species cluster together (Supplementary Fig. 3b), indicating that the genes duplicate before the speciation. In line with this, the inter-species sequence identity of the orthologous genes is higher than that of the intra-species paralogous genes (Supplementary Fig. 4). All duplicated gene copies were tandemly clustered at the trypsinogen gene locus, except the one that escaped by inter-chromosomal duplication around 30.9 Mya (95% CI: 26.6-35.0), which evolved into *PRSS3* (mesotrypsinogen) on chromosome 9 in the great ape lineage ^15^ (Supplementary Fig. 5). Followed by *PRSS3*, the ancestral copy of *PRSS2* was among the earliest duplication event at *PRSS1-PRSS2* locus in the great apes, which dates back to 29 Mya. A six-copy structure might have formed around 24 Mya, which became the major form at the *PRSS1-PRSS2* locus in non-human great apes. Collectively, these results indicate that this 6-copy structure is an ancestral form in the great ape lineage.

To further analyze how the ancestral 6-copy structure evolved into the human-specific deletion form, we aligned the short reads from great apes as well as from archaic and modern humans to the chimpanzee genome. The results revealed that the majority of great apes retained the 6-copy structure, while both archaic and modern humans lost the 1^st^ copy of the 6-copy structure (Supplementary Fig. 6a and 6b). Further sequence analysis showed that the loss was likely caused by non-allelic homologous recombination (NAHR), which resulted in a human-specific fusion form of 1^st^ (first 1.78-kb) and 2^nd^ (last 8.9-kb) copy structure (Supplementary Fig. 6d; Supplementary Note). However, the gene body of human *PRSS1* was not affected as it was mainly embedded in the 2^nd^ copy of the chimpanzee (Fig. 1a). We estimated that the fusion event started at 6.36 Mya (95% CI:5.12-7.71) after the split of human-chimpanzee, and fixed in the common ancestor of archaic and modern humans (Supplementary Fig. 6c). In the modern humans, the majority of the haplotype structures are the 3-copy or the 5-copy forms, with a very rare (0.1%, 4/4046) structure of 4-copy observed in the African population. In archaic hominids, both Altai and Vindijia Neanderthals have a 5-copy structure as a modern human, while Denisovan has a 4-copy structure with the homozygous deletion at the 5^th^ copy of chimpanzee 6-copy structure. Taken together, human trypsinogen genes experienced duplication expansion during Oligocene Epoch, and have decreased since divergence with the chimpanzee.

### Relaxed selection of human trypsinogen genes

Humans only retained *PRSS1* and *PRSS2* as protein-coding genes, while all six copies are protein-coding in non-human great apes. We hypothesize that the relaxation of natural selection facilitates accumulating nonsynonymous mutation which conducted the pseudogenized process at some of the duplicated genes. By calculating the non-synonymous change to synonymous change ratio (dN/dS), a measurement of natural selection pressure (Methods), we found higher dN/dS values at the 3^rd^, 4^th,^ and 5^th^ trypsinogen gene of the six-copy structure (corresponding to human *PRSS3P1, PRSS3P2*, and *TRY7*, respectively) than the other three genes, indicating the relaxation of selection pressure on these duplicated genes in the ape lineage (Fig. 1c; Supplementary Fig. 7a and 7b). Interestingly, these three copies were also absent in gibbon, suggesting that they are not functionally conservative (Supplementary Note and Supplementary Fig. 8). These results, together with the loss of the first protein-coding gene showing comparable dN/dS to *PRSS1* in humans, indicate that the trypsinogen genes might be functionally redundant during the evolution of apes.

When focused on the polymorphism of coding sequences in humans, we found that the dN/dS of *PRSS1* is comparable to that of the nonhuman great ape while the ratio of the *PRSS2* branch is larger than the paralog pseudogenes and significantly deviated from the background (P<0.01; Supplementary Fig. 7c), which indicates that the purifying selection is still acting on *PRSS1* but has been relaxed on *PRSS2*. Consistently, the non-synonymous mutations are not only more frequent and also associated with a larger impact on *PRSS2* than on *PRSS1* (Fig. 1d). These findings provide an evolutionary explanation of why the known pathogenic variants of the pancreatic disease were dominantly found at *PRSS1* but none were reported at *PRSS2* ^11^.

### Haplotype polymorphism in human populations

Taking advantage of deep-sequencing data, we characterized genetic diversity among the modern human populations. The proportion of 5-copy and 3-copy structures varies substantially among different populations (Supplementary Fig. 9a). In African populations, the 5-copy structure comprised more than 90% of the haplotypes. In non-African populations, the 3-copy structure is the major form (>70%) in East Asia and even fixed in Oceania, while the European populations carry both two forms with relatively more 5-copy (0.6-0.7) than 3-copy (0.3-0.4).

According to the single nucleotide variants at the *PRSS1-PRSS2* locus, we further classified the sequences of the modern human populations into 5 major groups (Fig. 2a and Supplementary Fig. 10a; Methods). The 5-copy structure (denoted as the long-form) shows a great diversity in all the major groups. Most of the long-form haplotypes were African-specific, especially for H3 and H5, while the long-form haplotype in the non-African haplotype is almost solely H2. In contrast to the large diversity, the 3-copy structure (denoted as the short-form) was dominantly H1S in worldwide populations and most (97.1%) of which were distributed in the non-African population. Overall, the haplotype structure in the non-African population is mainly (90.4%) comprised of the long-form H2 and the short-form H1S (Fig. 2b).

**Fig. 2.**
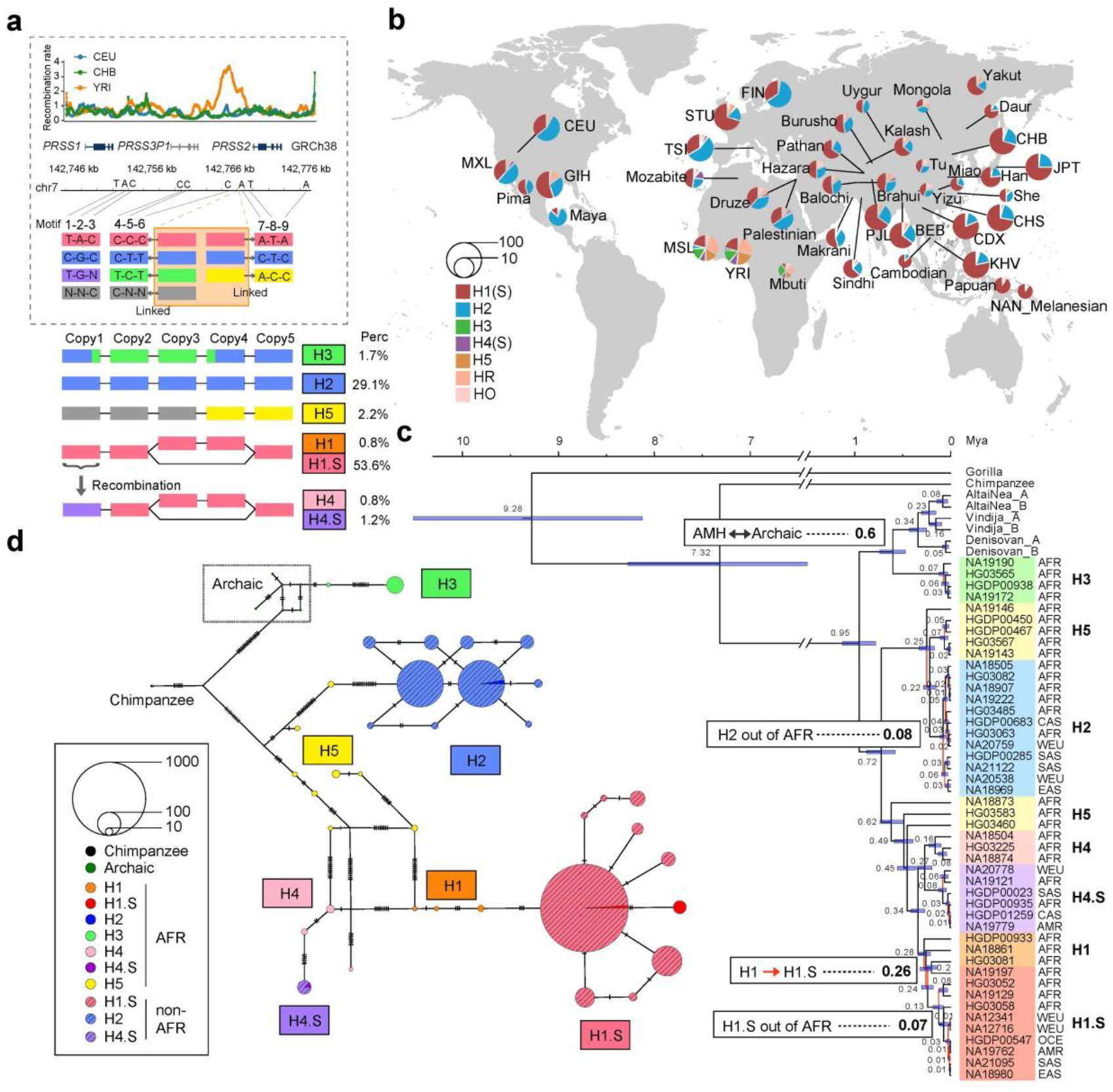
Haplotype diversity of trypsinogen genes in worldwide populations. **a**. Haplotype structure and recombination rate. The dash box shows the recombination rate (upper panel) and 3-nuclotide motif of each trypsinogen copy based on GRCh38 primary chromosome 7. The major haplotype structure is shown below the dash box with percentages in the 2,027 samples. **b**. Haplotype diversity distribution in the worldwide populations. **c**. Bayesian phylogenetic tree of major human haplotype groups. Blue bars show the 95% CI of divergence time. **d**. Network of the most common 20 major haplotypes in African and non-African populations, respectively. The phylogeny and network are based on the 3-copy sequence (*PRSS1-PRSS3P1-PRSS2*) shared between long-form and short-form haplotypes. The circle size in **b** and **d** indicates the sample size. AFR, African; WEU, West European; CAS, Central Asian Siberian; SAS, South Asian; EAS, East Asian; AMR, American; OCE, Oceanian.

The greater diversity of the long-form haplotypes in the African populations implies an African origin of a long-form structure. The topology of the phylogenetic tree based on regions (*PRSS1-PRSS3P1-PRSS2*) shared between long-form and short-form haplotypes show that the deepest divergence within the human lineage separates H3 and the archaic haplotypes from other modern human haplotypes ∼1 Mya (Fig. 2c). This divergence is caused by consecutive different alleles at *PRSS3P1* and *PRSS3P2* regions (Supplementary Fig. 11), which formed a typical Yin-yang haplotype structure ^16^. In the H3 (the Yin-lineage) branch, the archaic human lineage separated from African H3 samples ∼600 kya. Notably, we observed the African H3 haplotypes have a very short time to the most recent common ancestor (TMRCA) and a large extended haplotype homozygosity (EHH), indicating these Yin-lineage haplotypes are under recent positive selection (Supplementary Fig. 11). In the non-H3 (the Yang-lineage) branch, the African-specific H5 is the ancestral form of two major sub-lineages, i.e., H1/H4 and H2 groups (Fig. 2c). H2 was found in the African population and its divergence time with non-African H2 is ∼80 Kya. Notably, a previously reported pancreatitis-protective mutation G191R ^17^ is exclusively located on H2 (Fig. 1d). The H1 group has a deeper African-specific lineage (H5-1/H5-4) and was involved in the formation of many other haplotypes via recombination (Supplementary Fig. 10b and 10c). The short-form H1S was derived from H1 around 260kya before out of Africa. Notably, by analyzing the sequence breakpoint and divergence, we concluded that other haplotypes (H4S and HR3S) with the same deletion of *PRSS3P2* and *TRY7* originated from H1S via recombination (Supplementary Fig. 10c and Supplementary Note). The divergence time between African and non-African H1S was ∼70 kya. Further network analysis confirmed the African origin and the relationship of the major haplotypes (Fig. 2d). Taken together, the major non-African haplotypes have an African origin and belong to the Yang-lineage with long-form as the ancestral state (Fig. 2c).

### Human *PRSS2* on the short-form haplotype is under positive selection

On the amino acid level, we applied the dN/dS ratio to test natural selection for each trypsinogen gene copy on two haplotype structures in different populations. In contrast to a strong purifying selection on *PRSS1*, the selection pressure is reduced on *PRSS2* in non-African populations for both long and short-form haplotypes. Especially in the short form, the overall populations and the East Asian branches have a signature of positive selection on *PRSS2* (dN/dS>1; P<0.05, likelihood ratio test; Fig. 3a and Supplementary Fig. 7d).

**Fig. 3.**
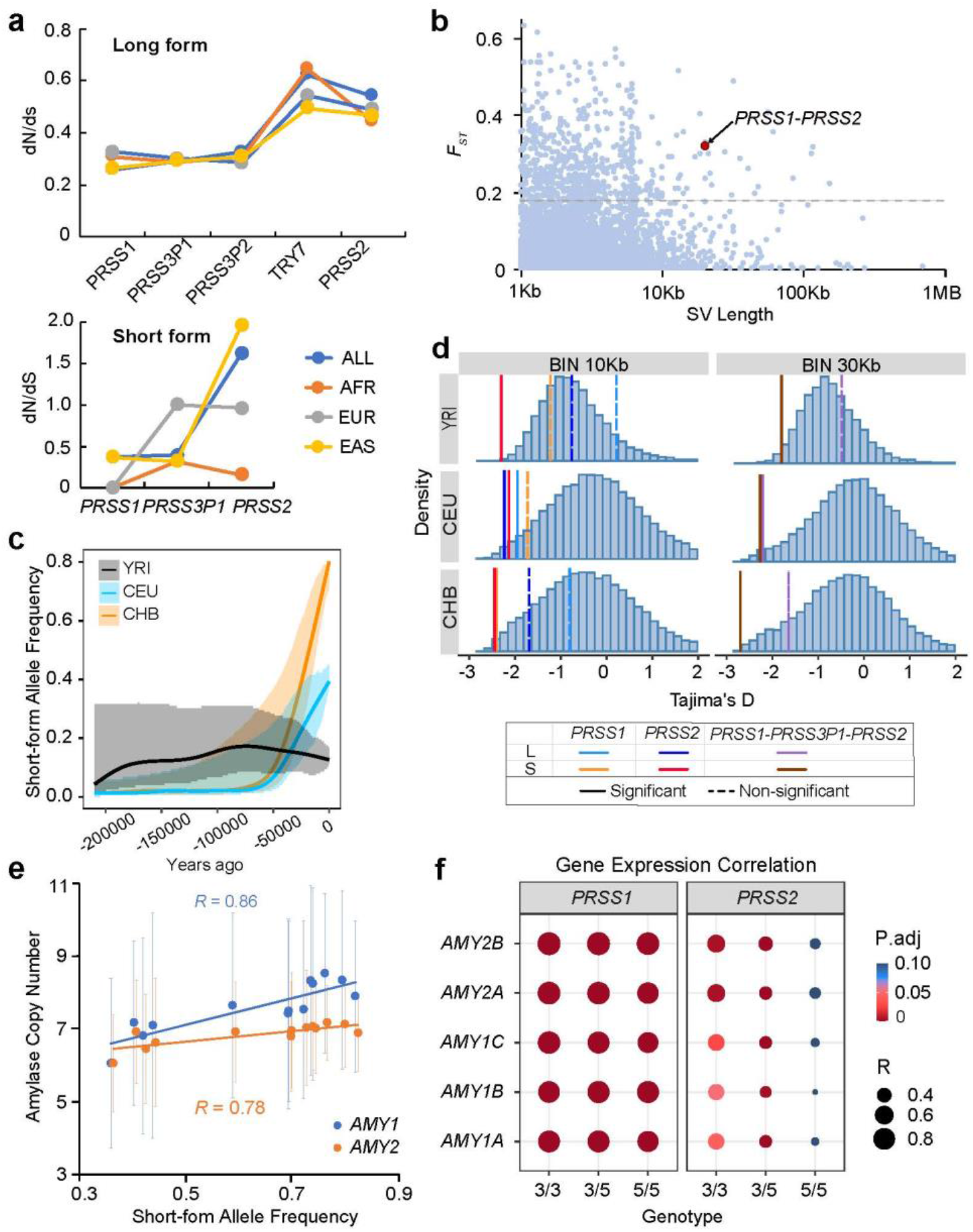
Positive selection on *PRSS2* of human short-form haplotype. **a**. dN/dS ratio for different trypsinogen gene copies in diverse populations. See phylogeny of the branch test in Supplementary Fig. 7c and 7d. **b**. Distribution of cross-population F_ST_ (YRI-CEU-CHB) for genome-wide structural variants (SV) with length >1kb. Each dot denotes an SV and the red circle represents the structural polymorphism at *PRSS1-PRSS2* locus. Dash line indicates the 5% cut-off of F_ST_ under demographic simulation. **c**. Inferred allele frequency trajectory of short-form haplotype. **d**. Tajima’s D distribution of 10-kb and 30-kb sequences on chromosome 7. The vertical lines denote the corresponding values at the *PRSS1-PRSS2* locus. **e**. Correlation of average amylase gene copy and short-form frequency of trypsinogen gene haplotype in non-African populations. The bars denote the standard deviation of amylase gene copy number in each population. **f**. Gene expression correlation between the amylase genes and trypsinogen genes (*PRSS1* and *PRSS2*). The number in the genotype indicates the haplotype structure: 3, short-form; 5, long-form.

On the frequency level, this structural polymorphism is among the top stratified structural variants (SVs) among African, European, and East Asian populations, and is less likely explained by demographic events (empirical P-value<0.05; Fig. 3b; Supplementary Fig. 9b; Methods), which suggests that natural selection might account for the substantial variation. Using a coalescence approach (Methods), we reconstructed the frequency trajectory of the short-form haplotype (Fig. 3c). Consistent with our phylogenetic analysis (Fig. 2d), the short-form haplotype spread out of Africa 70-80 kya. Positive selection started in East Asian 37 Kya and the coefficient (*s*) is estimated to be 0.0023 (log-likelihood ratio=3.1), while the selection is weaker in European (*s*∼0.0012; log-likelihood ratio=1.6). The selection might be still ongoing in the Eurasian populations, evidenced by a star-like pattern of the network in the short-form haplotypes but not in the long-form haplotype (Supplementary Fig. 15), suggesting a recent selective sweep. Furthermore, Tajima’s D of the short-form haplotype showed significantly negative values compared to both the background with genomic windows and simulation of the demographic model (Fig. 3d and Supplementary Fig. 16; Methods), which confirms the signal of a selective sweep in the Eurasian populations. Collectively, these results consistently indicate that the short-form haplotype is under positive selection.

Since trypsin plays an essential role in digesting proteins, diet might be one of the selection driving forces. A previous study showed the positive selection acting on the copy number of salivary amylase gene in the populations with high-starch intake ^18^, here we observed a strong positive correlation between the average copy number of both salivary and pancreatic amylase genes (i.e., *AMY1* and *AMY2*, respectively) and the short-form frequency in the non-African populations (R>0.78; Fig. 3e). By comparing the correlations of amylase genes with other randomly selected loci, we found this high interchromosomal correlation is less likely explained by chance (P<0.05; Supplementary Fig. 17a). Furthermore, at the transcription level, we found a strong expression correlation between amylase genes and trypsinogen genes, especially for *AMY2* and *PRSS1* (Supplementary Fig. 17d and Fig. 3f). The pancreatic co-expression network constructed by weighted correlation network analysis (WGCNA) ^19^ revealed that the amylase and trypsinogen genes were clustered in the same module with other genes enriched in pancreatic secretion, in which *AMY2B* and *PRSS1* together with the pancreatic lipase gene *PNLIP* are the hub genes (Supplementary Fig. 17b-e). The co-expression is strong (R>0.8) between *PRSS1* and amylase genes regardless of the *PRSS1-PRSS2* haplotype, but it varies between *PRSS2* and amylase genes across different configurations (Fig. 3f). Particularly, the homologous short-form haplotypes show significantly positive-correlated expression between *PRSS2* and *AMY1* genes (R>0.45; adjusted P<0.05) while no such significant correlation found in the homologous long-form samples (adjusted P>0.1). Collectively, both genomic and transcriptomic results consistently indicate that diet might simultaneously drive the evolution of amylase and trypsinogen genes, in which the short-form *PRSS1-PRSS2* haplotype leads to a more orchestrated expression pattern for the two types of digestive enzymes in the pancreas tissue.

### Pseudogenes exert as tissue-specific enhancers for *PRSS2* expression in the pancreas

The Genotype-Tissue Expression (GTEx) data from multiple tissues show that *PRSS2* is tissue-specific and the top expressed gene in the pancreas ^20^ (Supplementary Fig. 18). The copy number of trypsinogen genes at *PRSS1-PRSS2* locus has a significant positive-correlation with *PRSS2* expression level, i.e., the 5-copy haplotype is associated with higher *PRSS2* expression compared with the 3-copy haplotype (Fig. 4a). Notably, the haplotype structure is in complete linkage with SNV rs2855983, an expression quantitative trait loci (eQTL) variant with the largest effect size among all the *PRSS2*-eQTL variants in the pancreas tissue (Fig. 4b). Moreover, the *PRSS2* eQTL variants with stronger LD of rs2855983 (R^2^>0.9) tend to have a larger effect size than the rest eQTL variants (P=6.7e-05; Supplementary Fig. 19a). Consistently, a strong allelic specific expression (ASE) effect was observed for *PRSS2* in the 3-copy/5-copy heterozygous carriers, in which the 5-copy haplotype tends to produce more mRNA than the 3-copy haplotype (P<0.0001; Supplementary Fig. 19b). These results suggest that the copy number variation of the pseudogenes is a lead genetic factor among the common genetic variants (minor allele frequency, MAF>0.1) accounting for *PRSS2* expression change in pancreas. Furthermore, we observed that the pancreatic co-expression between *PRSS1* and *PRSS2* is significantly different in diverse haplotype configurations (P<0.001; Fig. 4a), with the highest correlation coefficient in the homologous 3-copy haplotype carriers (0.778) and the lowest in the homologous 5-copy carriers (0.474). Interestingly, albeit in low expression in other tissues, these two genes were highly correlated (R>0.8) regardless of the haplotype configurations (Fig. 4c and Supplementary Fig. 19c). These observations indicate that the expression of the two genes is synchronized in most of the tissues while disrupted by the long-form haplotype in the pancreas.

**Fig. 4.**
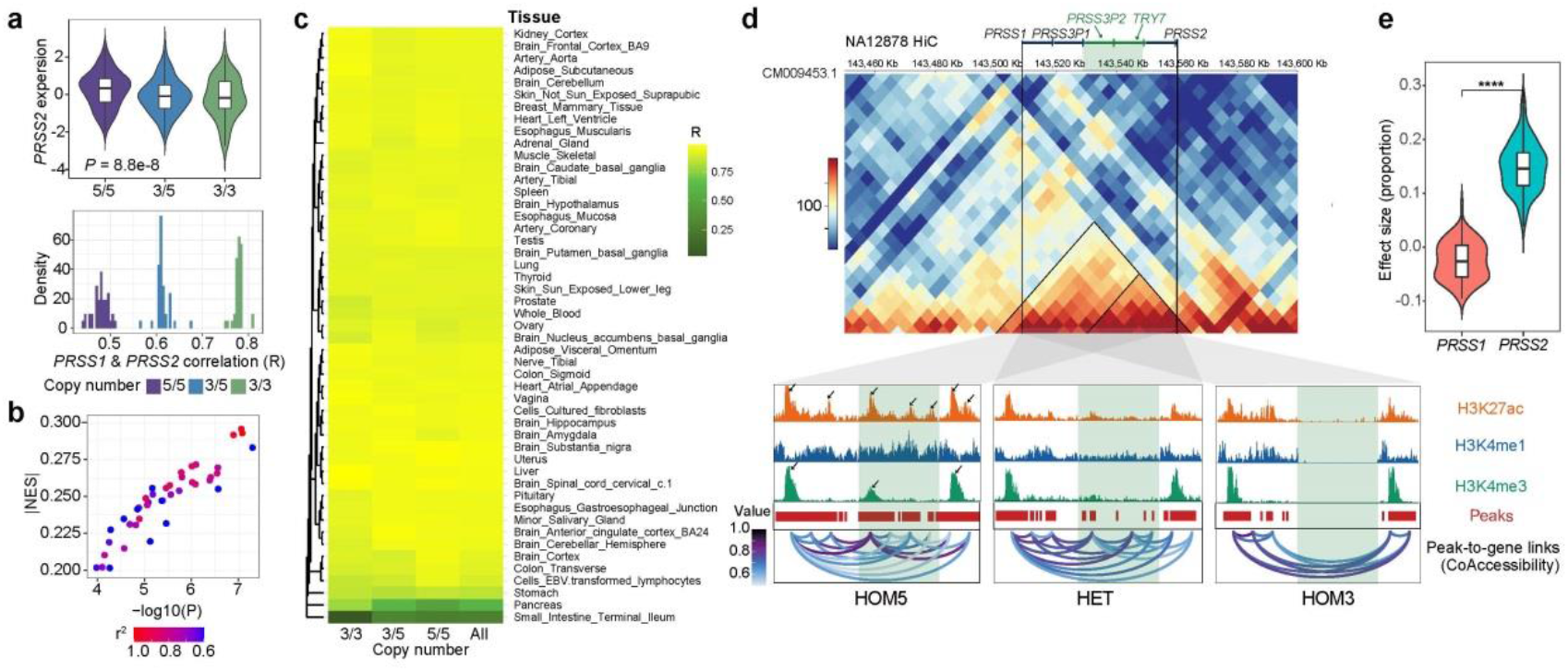
Trypsinogen pseudogenes increase *PRSS2* expression in pancreas. **a**. The copy number structure is associated with *PRSS2* expression (upper panel) and affects the co-expression between *PRSS1* and *PRSS2* (lower panel). **b**. Distribution of normalized effect size (NES) and eQTL P-values for *PRSS2* eSNVs. Each circle represents an eSNV and the color indicates the LDs with the haplotype structure. **c**. Correlation coefficient of gene expression between *PRSS1* and *PRSS2* in different haplotype configurations across multiple tissues. **d**. Integration of epigenetic signals at *PRSS1-PRSS2* locus from omics data. Upper panel, a Hi-C contact map from GM12878. Black lines indicate the TADs. Lower panel, pancreatic histone modification and coaccessibility of open chromatin signals in different haplotype carriers. The histone modification signals are derived from ChiP-seq and the loops in the coacessbility are inferred from scATAC-seq. HOM5, homologous long-form; HOM3, homologous short-form; HET, heterozygous long-form/short-form carriers. The shaded green denotes the polymorphic pseudogenes. In this analysis, sequence from a *de novo* assembly (NA12878) that has a homologous long-form form is used as reference genome in this analysis. **e**. Comparison of effect size of copy number structure between *PRSS1* and *PRSS2* in terms of proportion of expression level change. The violin plot was based on randomly sampling 100 samples.

Given that the two pseudogenes *PRSS3P2* and *TRY7* are the major difference between the long-form and short-form haplotype, we hypothesized that the pseudogene enhancers as a legacy of duplication events were still functional in the pancreas, although their gene bodies no longer produce protein. To test this, we first investigated the topological associated domain (TAD) with Hi-C data by using a homologous short-form and a homologous long-form assembly as reference (Methods). The contact map of the short-form structure shows a large TAD for *PRSS1, PRSS3P1*, and *PRSS2* and a nested one in *PRSS3P1* and *PRSS2* (Supplementary Fig. 19d). For the 5-copy structure, all five trypsinogen gene copies are in a large TAD and the *PRSS2* is in a sub-TAD with *PRSS3P2, TRY7* (Fig. 4d). These contact maps suggest of the physical interaction between *PRSS2* and the pseudogenes. To explore whether the homologous sequences (*PRSS3P2* and *TRY7*) show signatures of enhancers, we analyzed the histone modifications with Chromatin Immunoprecipitation sequencing (ChiP-seq) data from pancreas tissue (Methods). In addition to the typical enhancer signals at the promoter regions of *PRSS1* and *PRSS2*, we also observed strong peaks of H3K27ac and H3K4me1 at the *PRSS3P2* and the *TRY7* regions in the homologous 5-copy samples, which indicates these sequences at the pseudogene copies might still exert their enhancer functions. Concordantly, these peaks were weaker in the heterozygous 3/5-copy sample and absent in the homologous 3-copy sample (Fig. 4d).

To further link these enhancer signals to the target genes, we leveraged multi-omic single-cell sequencing data of Assay from Transposase-Accessible Chromatin with high-throughput sequencing (ATAC-seq) and RNA sequencing (RNA-seq), which can simultaneously detect the epigenomic state and measure the transcription level at the same cell. In pancreas tissue, we integrated the peak accessibility and gene expression from the same cells and identified the loops between the polymorphic pseudogenes (*PRSS3P2* and *TRY7*) and the two protein-coding genes (*PRSS1* and *PRSS2*) in the long-form haplotype carrier (Fig. 4d and Supplementary Fig. 20a). The inferred coaccessibility indicates that the chromatin states of these trypsinogen genes are co-activated simultaneously in pancreas cells and that *PRSS1* and *PRSS2* are potentially regulated by the pseudogene enhancers at the single-cell level. We further analyzed the bulk RNA-seq data to quantitatively assess the regulatory impact on *PRSS1* and *PRSS2*. The eQTL results showed that these pseudogene enhancers are significantly associated with an increase of *PRSS2* expression in

∼14,500 Transcripts Per Million (TPM), which corresponds to a mean of ∼15% gene expression level per haploid; however, no significant association was observed for *PRSS1* expression (Fig. 4e). Consistently, the expressions of *PRSS1* and *PRSS2* showed no significant difference for the homologous short-form samples (P=0.503, t-test), in which the polymorphic pseudogenes are absent; however, the expression of *PRSS2* is significantly higher than that of *PRSS1* for the homologous long-form samples (P<2.0e-8, t-test), in which the polymorphic pseudogenes are present (Supplementary Fig. 19c). Furthermore, we found that the H3K27ac peaks at the polymorphic pseudogenes (*PRSS3P2* and *TRY7*) had higher sequence similarity with the known *PRSS2* enhancers, which potentially provided the transcription binding sites that are specific to motifs of *PRSS3P2, TRY7* and *PRSS2* but absent in *PRSS1* (Methods; Supplementary Fig. 19e). These lines of evidence suggest that the trypsinogen genes at *PRSS1-PRSS2* locus are activated synchronously and the polymorphic pseudogene enhancers might specifically upregulate *PRSS2* expression in pancreas.

Noted that the *PRSS1*-*PRSS2* co-expression was reduced in the long-form haplotype in the pancreas but not in other tissues (Fig. 4c and Supplementary Fig. 19c), we hypothesized that the trypsinogen pseudogene enhancers are specifically activated in the pancreas. To test this, we examined the co-accessibility of the trypsinogen genes from long-form haplotype carriers in other tissues. In the spleen tissue of a homologous long-form sample, we observed the open chromatin peaks only present at *PRSS1, PRSS3P1*, and *PRSS2* promoter regions, but almost absent at *PRSS3P2* and *TRY7* regions (Supplementary Fig. 20a). Furthermore, we found much lower signals of H3K4me1 histone modifications on *PRSS3P2* and *TRY7* in the spleen than those in the pancreas for the same samples, indicating these enhancers were inactivated in the spleen (Supplementary Fig. 20b). Given the large difference of *PRSS1*-*PRSS2* co-expression patterns for long-form haplotype carrier between the pancreas and other tissues (Fig. 4c), these results not only suggest of a tissue-specific manner of the polymorphic pseudogene enhancers, but also supported that the activation of pseudogene enhancers might increase *PRSS2* expression and disrupt *PRSS1-PRSS2* co-expression.

### Pseudogenes contribute to pancreatitis risk as a causal factor

To further explore the biomedical implications of the haplotype structure, we searched the SNVs in complete LD with the haplotype structure in the genome-wide association study (GWAS) catalog. Most strikingly, rs2855983, the lead *PRSS2*-eQTL SNV with the complete linkage of the haplotype structure, is the top SNV associated with alcoholic chronic pancreatitis (P=1e-40) in a previous large-scale association study of the European population ^21^. We performed a meta-analysis based on the LD information and estimated the odds ratio (OR) for a long-form haplotype structure to chronic pancreatitis is 1.79 (Supplementary Fig. 21), consistent with the finding that elevated *PRSS2* expression is detrimental in pancreatitis ^22^. Furthermore, we observed a significant association between the haplotype structure and the expression ratio of *PRSS1*/*PRSS2*, with the highest ratio in homologous 3-copy haplotype carriers and the lowest ratio in homologous 5-copy haplotype carriers (Supplementary Fig. 22a), which agrees with the previous medical observation that pancreatitis patients have lower or even reverted ratio of cationic and anionic trypsinogens (*PRSS1/PRSS2*) than the controls ^23^.

Still, it is unclear about the causal relationship between this ratio and pancreatitis, neither is the causal factor accounting for the common genetic variant associated with pancreatitis risk ^12^. Taking advantage of GWAS summary statistics and the gene expression data, we applied Mendelian Randomization methods to evaluate the causal relationship between the *PRSS1*/*PRSS2* ratio and the disease. The Summary-based Mendelian Randomization (SMR) results support a causal role in pancreatitis (Fig. 5a), and the pancreatitis risk is mediated by elevated *PRSS2* expression while not by *PRSS1* expression (Supplementary Fig. 22b and 22c). Utilizing a colocalization approach, we further identified the polymorphic pseudogenes as a single causal variant accounting for the gene expression ratio and the pancreatitis risk (posterior probability>0.99) rather than previously identified GWAS variants (Fig. 5a). Collectively, these results indicate that the polymorphic pseudogenes are a causal factor of the common variants contributing to the susceptibility of pancreatitis by mediating *PRSS2* expression.

**Fig. 5.**
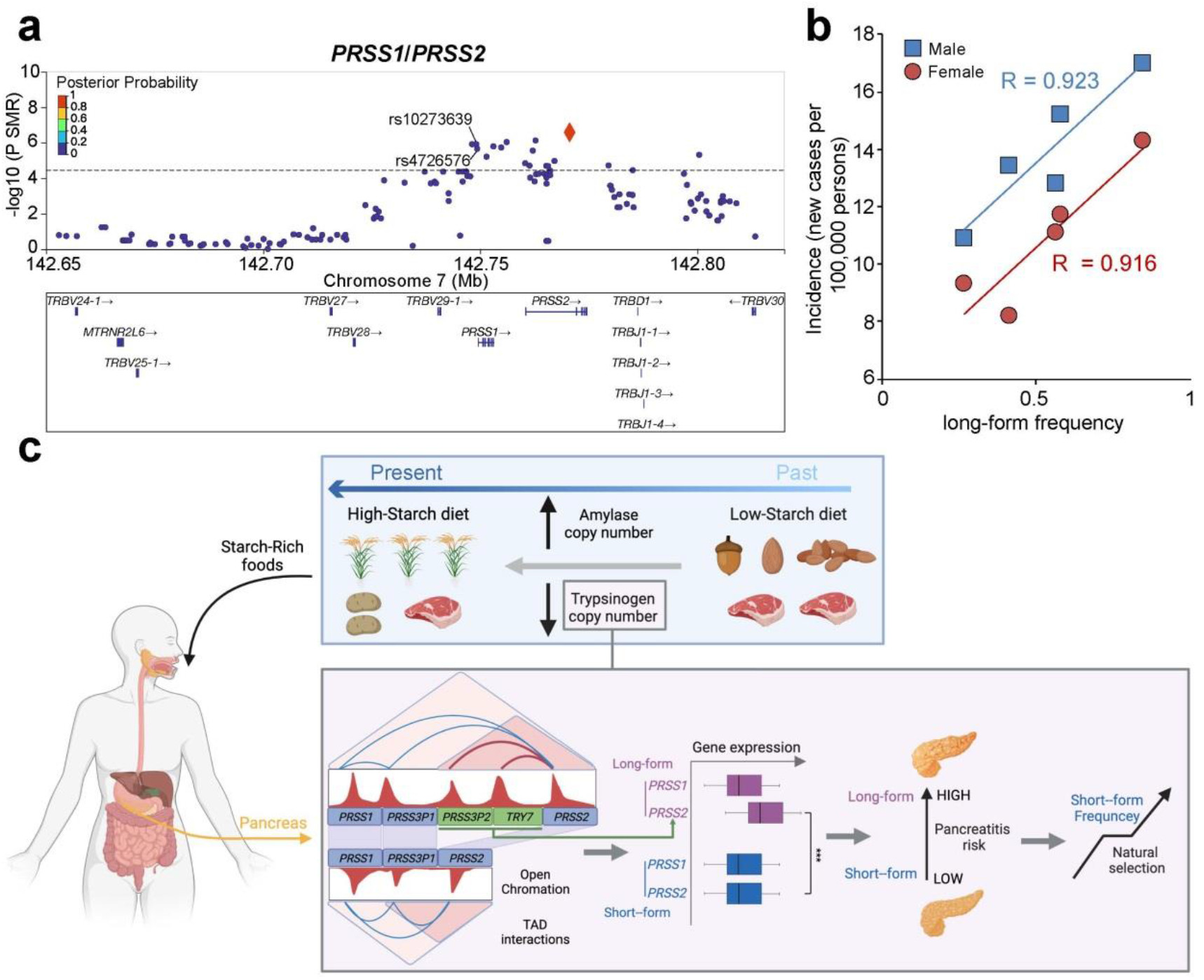
Trypsinogen pseudogenes confer risks to human pancreas disease. **a**. Summary-based Mendelian Randomization (SMR) and colocalization analysis using *PRSS1*/*PRSS2* expression ratio and GWAS summary statistics identified the pseudogenes as a single causal factor of common variants contributing to pancreatitis. The y-axis shows P-value of SMR and the dash line denotes the multiple-testing cut-off. Each circle dot represents a genetic variant, and the diamond denotes the tagged variant (rs2855983) of complete LD with haplotype structure. The filled colors of the diamond and the circle indicate the posterior probability of being a shared single causal variant accounting for *PRSS1*/*PRSS2* expression ratio and odds ratio of pancreatitis. The SNVs marked with IDs were the previously identified GWAS variants. **b**. Correlation between incidence of pancreas cancer and long-from haplotype frequency across different ethnic groups in the United States. **c**. A model of trypsinogen gene evolution in human. When digesting food, both the amylase and trypsin would be simultaneously activated in pancreas due to their tightly co-expressed pattern. The high-starch food requires more amylase which in turn produces more trypsin. Compared with the long-form trypsinogen haplotype, the short-form haplotype without the pseudogene enhancers could significantly reduce *PRSS2* expression, which protects pancreatic cells from autodigestion and lowers pancreatitis risk. Such trait accompanied with higher copy of salivary amylase genes is favored by natural selection when diet shifts to high-starch food.

In addition to pancreatitis, *PRSS2* was found to be a biomarker of pancreas cancer for its elevated expression in patients ^24^. Using pancreas cancer statistics from the Surveillance, Epidemiology, and End Results (SEER) Program, we observed a strong correlation between the frequency of long-form haplotype and the incidence of pancreas cancer in different ethnic groups in the United States (R2>0.9; P<0.01; Fig. 5b; Methods), which suggests the common long-form haplotypes might also be a risk factor contributing pancreas cancer risk in the general populations.

The higher risk of the long-form haplotype is likely explained by diet shifts in an evolutionary perspective (Fig. 5c): Based on the report that pure glucose meal could stimulate a submaximal trypsin secretion similar to meal containing proteins ^25,26^ and the co-expression nature of amylase and cationic trypsin (Supplementary Fig. 17), high-starch diet might lead to high *PRSS1* expression in pancreas. The stimulation of the cationic trypsinogen gene simultaneously activates other trypsinogen genes including the pseudogene enhancers (Fig. 4d and Supplementary Fig. 20). As a consequence, one long-form haplotype generates ∼15% more anionic trypsin than the short-form haplotype (∼30% for homologous diploid) in the pancreas. The excessive secretion of trypsin could increase the risk of proteases causing damage to the pancreatic acinar cells ^27^, especially when the glucose proportion of nutrition ingredients increases during a high-starch diet shift. Therefore, the short-form *PRSS1-PRSS2* haplotype associated with lower anionic trypsinogen expression is more protective to the pancreas and more adaptive for agricultural societies.

## Discussion

Gene duplication plays an important role in evolution, which can generate diverse biological consequences for the new copy, such as keeping the original function and evolving novel function ^28^. Human trypsinogen genes are a typical product of gene duplication, which emerged before the speciation of apes during the Oligocene Epoch when mammals increased in numbers, diversity, and size. The emergence of duplicated trypsinogen genes might have helped the ancestries of apes better absorb protein during diet change when the woodlands were replaced by grassland expansion at that time ^29^, but the loss of trypsinogen genes tends to be more favored by natural selection after the speciation of apes, as evidenced by gibbon and humans (Fig. 1b). The former is frugivore and has only three copies, while the latter has a much diverse polymorphism with only two protein-coding genes. In addition, the loss of copy polymorphism is also found in chimpanzees, but none of the copy number gains is observed in either the general human population or the non-human great apes (Supplementary Fig. 6a). On the contrary, copy number gain of the trypsinogen genes in human were found to be detrimental and causes hereditary pancreatitis ^9,30^. These observations all indicate the redundancy of duplicated trypsinogen genes in the human lineage.

High-starch diet shift is likely to account for the positive selection of the short-form haplotype favoring lower trypsin traits, supported by the unexpected high correlation between amylase and trypsinogen genes at both genomic and transcriptomic levels. This in turn suggests that the adaptation to diet shifts is a complex process that involves multiple genes as the targets of natural selection. Particularly, the short-form haplotypes are fixed in the Papuan population (Supplementary Fig. 9b), whose diet is mostly vegetarian. In addition to a high-starch diet, trypsin inhibitor foods could be another driving force of positive selection in favor of the short-form haplotype. Trypsin inhibitors are found in many food plants and function as a natural defense system against herbivores ^31^. Archeological evidence showed that starch grains, beans, and yam have been human food resources since the Paleolithic period ^32,33^. These plants are the well-known natural source of trypsin inhibitor which has an antinutritional effect on vertebrates ^34^. Rats fed with soybean trypsin inhibitor would cause hypertrophy of the pancreas ^35^. Although the trypsin inhibitor toxicity to humans is still unknown, it is clear that consuming trypsin inhibitor stimulates the secretion of human trypsin as a feedback regulation *in vivo* ^36^. This process increases the risk of pancreas disease if excessive trypsin products were secreted. Because the short-form structure enables a significant lowering of the anionic trypsinogen expression for an average of 15-30% compared with the long-form structure for a diploid genome, the risk of pancreatitis can be reduced when the feedback regulation is intrigued with simultaneous activation of trypsinogen genes followed by consuming trypsin inhibitor foods.

In summary, the evolution of trypsinogen genes provides an example of how diet change shapes their genetic diversity jointly with the salivary amylase genes ^18^, and a model of how the redundancy of gene duplications contributes to the risk of human disease.

## Methods

### Evolution history analysis of trypsinogen genes in the primates

To estimate the number of trypsinogen genes in the primates, we searched for the alignments with BLAST by using the human trypsinogen copy sequence (chr7_KI270803v1_alt:749409-801557) as a query. The non-human primates analyzed here included the sequences from Mouse lemur, Tarsier, Ma’s night monkey, Capuchin, Macaque, Gelada, Black snub-nosed monkey, Sooty mangabey, Olive baboon, Gibbon, Orangutan, Gorilla, Chimpanzee, and Bonobo. A typical trypsinogen gene copy unit including genic and intergenic regions is 10.6-kb. For the trypsinogen gene clusters, we named the duplicated copies by order in the forward strand. We applied Mummer (v3.23) ^37^ and lastz (v1.04) ^38^ to calculate the sequence identity of different duplication copies for different primates. Miropeats (v2.02) ^39^ was used to visualize the sequence structure between different primates. The species divergence time was derived from TimeTree ^40^.

To explore the evolution of different duplication copies in primates (Supplementary Table 1), we first aligned the sequences of each copy with MAFFT (v7.310) ^41^, and constructed the phylogenetic tree with BEAST (v2.6.7) ^42^. We used the HKY with GAMMA category count=5 for the site model. We applied the strict clock for the Clock model and the Calibrated Yule Model for tree priors. Gamma (0.001, 1000) was used as the prior distribution of clock rate and the divergence time of capuchin-ma’s night monkey was utilized as the calibration following a log-normal (M=18.3, S=0.05) distribution. The MCMC chain length was set to 1,000,000, and the first 10% runs were used as burn-in. We used Figtree (v1.4.4) to visualize the phylogenetic tree.

To estimate the age of the NAHR event from the ancestral 6-copy haplotype to the human 5-copy haplotype, we performed BEAST analysis with the first trypsinogen gene copy (*PRSS1*) of archaic and modern humans together with manually fused sequences from the first 1.7kb and the second 8.9kb of nonhuman great ape’s gene copy. The divergence time of orangutan-human was set as the calibration following a log-normal (M=16, S=0.09) distribution. Other parameters were the same as above.

To estimate the selection pressure for different trypsinogen genes, we applied the maximum likelihood approach to estimate the non-synonymous to synonymous mutation ratio (dN/dS) in primates by using PAML (v4.9j) ^43^. We used a branch model to calculate the evolving rate of each copy with a constant rate in a null model (model=0) and with a lineage-specific rate in an alternative model (model=2). Likelihood ratio tests (LRT) for each copy in two models were compared with the chi-square test (df=1). To investigate whether there are sites under selection, we performed a site model analysis for the branches with two protein-coding genes (*PRSS1* and *PRSS2*). Only transcripts with the conserved primate trypsinogen gene sequence (247 amino acids) were considered in this model. We performed both the M1a-M2a comparison and the M7-M8 comparison with the Bayes empirical Bayes method.

### Short-read alignment and variant calling

To estimate the copy number of trypsinogen genes at the *PRSS1-PRSS2* locus in the great ape lineage, we aligned the short sequencing reads of 25 chimpanzee, 13 Bonobo, 31 gorilla, and 10 orangutan samples from a previous study ^44^ to a chimpanzee assembly (GCA_002880755.3) ^45^. The short reads were mapped with aligner BWA (v0.7.17) ^46^ and the copy number was estimated based on the read-depth of the alignments with SAMtools (v1.6) ^47^ and adjusted by the depth of *PRSS1* and *PRSS2*.

To detect the genetic variants segregating in the human diverse populations at the *PRSS1-PRSS2* locus, we included 1,430 samples from 1,000 Genomes Project (KGP) ^48^ and 597 samples from Human Genome Diversity Project (HGDP) ^49^, both of which were sequenced with high coverage short-reads (∼30×). The detailed sample information was listed in Supplementary Table 2. The short sequencing reads were firstly aligned to the human reference genome GRCh38 with BWA, and then remapped to the GRCh38 alternative contig (chr7_KI270803v1_alt) to call the genetic variants including SNVs, small insertions, and deletions (INDEL) and copy number states with NGS.PRSS1-2caller (v1.5) ^50^.

### Characterization of haplotype in human population

To determine the haplotypes, we applied Beagle (v3.2.2) ^51^ to phase the genetic variants detected in 2,027 KGP and HGDP samples. We defined the haplotypes with a 5-copy structure as long-form, and the others with less than five copies (i.e., 3 or 4 copies) as short-form. Based on the loci with MAF>1% shared between long- and short-form haplotypes, we employed the unweighted pair group method with arithmetic mean (UPGMA) to cluster the haplotypes into groups. The clustering tree showed Three major branches (Supplementary Fig. 10a). One major branch we defined as the H3 group, which formed a typical Yin-Yang haplotype pattern ^16^ at the *PRSS3P1* and *PRSS3P2* regions, and its haplotype pattern could be easily distinguished from other groups (Supplementary Fig. 11), the rare 4-copy haplotype (H3S) also belongs to this branch. Another major branch, which contains the long-form haplotype dominantly (>90%) found in non-African populations, was named H2. The remaining branch is a large cluster that was further divided into three major groups (H1, H4, and H5). In this cluster, H1 and H4 are clustered in one branch. Their haplotypes showed differences in the first copy but their last four copies were similar. Both H1 and H4 had a deletion form of a 3-copy structure such as the primary scaffold in GRCh38, which was named H1S and H4S, respectively. Another branch in this cluster is the H5 group, which has a unique allelic configuration in the last two copies and is exclusively found in the African population.

To investigate the recombination rate at the *PRSS1-PRSS2* locus, we employed LDhat (v2.2a)^52^ to make inferences from African, European, and East Asian populations, respectively. The pattern of inferred recombination rate is consistent with that of the LD, in which African have strong LDs between the 2^nd^ and 3^rd^ copy and between the 4^th^ and 5^th^ copy. Based on these observations, we further selected the 3-SNV motif to distinguish different haplotypes (Supplementary Table 3). All the major haplotypes (H1 to H5), which comprises ∼91% of all the haplotypes, could be well-tagged by the three 3-SNV motifs. For the remaining haplotypes (7.2%) showing typical recombination, we could also use the combination of the motifs to characterize these sequences. Taken together, ∼99% of the haplotypes can be well captured by these 3-SNV motifs.

### Haplotype evolution history inference

The phylogenetic tree of both long-form and short-form haplotypes in worldwide populations was constructed with BEAST. The parameters were the same as the estimation of human fused trypsinogen gene copy except that the divergence time of chimpanzee-human (M=6.6, S=0.09) and gorilla-human (M=10.5, S=0.09) was used for calibration. The sequences shared by long-form and short-form, i.e., the 1^st^, 2^nd^, and 5^th^ human copy, were used in the phylogeny construction. We further conducted network analysis to explore the relationship of the major haplotype groups with PopART (v1.7) ^53^. For the haplotype network in the worldwide population, we used the most common 20 haplotypes of African and non-African populations, respectively, which comprised 79% of the total haplotypes. The recombined haplotypes were not included. For the haplotype network in the Eurasian populations, all haplotypes in Han Chinese in Beijing (CHB), and Utah residents with European ancestry (CEU) were included.

To investigate how the long-form haplotypes evolved, we constructed the phylogeny based on the long-form haplotypes (all five copies included) in the African population by using BEAST with the same parameters above. In addition, we calculated the TMRCA of each haplotype group and the pairwise haplotype group divergence time as ref ^54^. The inference of evolutionary history was based on the phylogenetic tree and the time (TMRCA and divergence time) estimation.

### Natural selection analysis in human populations

To test whether the positive selection is acting on trypsinogen genes in the human lineage, we constructed the phylogenetic tree using minimum evolution with coding sequences of both long and short-form haplotypes in African, European, and East Asian populations. The amino acid changes in 2,027 KGP and HGDP samples with respect to GRCh38 alternative contig were listed in Supplementary Table 4. Based on this phylogenetic tree, we used PAML to perform a branch test (Model 2) by specifying the *PRSS1* and *PRSS2* as a foreground branch, as well as for long form and short form haplotypes in each population, respectively. The likelihood ratio test was used to test the significance of positive selection for the branch with a dN/dS ratio>1 in Model 2 against the null model with the same dN/dS ratio.

We further applied three different approaches to detect natural selection in the African, European, and East Asian populations. Yoruba in Ibadan (YRI), CEU, and CHB from the 1,000 genomes project were used in this analysis. In the population-stratified approach, we characterized the differentiation of haplotype structure (long-form and short-form) in diverse human populations with F_ST_ ^55^. The empirical P-value was based on the background SVs which were documented in a comprehensive call-set generated by *de novo* assemblies ^56^. In the haplotype diversity approach, we calculated the statistic Tajima’s D for each duplicated copy of trypsinogen genes in both long-form and short-form haplotypes for shared regions (*PRSS1-PRSS3P1-PRSS2*), and compared the statistic with those calculated from 10-kb as well as a 30-kb sliding window on chromosome 7 to generate an empirical P-value. Given that the demographic events might affect the above two statistics, we also performed a simulation analysis with the typical three population out-of-Africa model by using stdpopsim (v0.1.2) ^57^ as a neutral scenario with effects from demographic events. The demographic parameters were set as described in a previous study ^58^. The observed F_ST_ and Tajima’s D were further compared with the simulations that matched the local feature at the *PRSS1-PRSS2* locus. We used R package rehh (v2.0) ^59^ to perform extended haplotype homozygosity (EHH) analysis for the H3 haplotype. In the coalescence approach, we applied to RELATE (v1.0.8) ^60^ and CLUES (v1) ^61^ to infer the allele frequency trajectory of the short-form haplotype over time and estimated the selection coefficient. We focused on the A-allele frequency trajectory of SNV rs2855983, which is employed as a tag of the short-form haplotype (R^2^=0.96, 1, for YRI, CEU, CHB, respectively). The mutation rate was set to 1.25×10^−8^ per base per generation.

### Correlation analysis between amylase and trypsinogen genes

At the genomic level, we calculated the correlation between the average number of *AMY1* and *AMY2* and the frequency of short-form haplotypes in 13 non-African populations from the 1,000 Genomes Project (Supplementary Table 2). The copy number of amylase genes (*AMY1* and *AMY2*) were genotyped by CNVnator (v0.3.3) ^62^. *AMY1* copy number was determined by the sum of *AMY1A, AMY1B*, and *AMY1C*, and the *AMY2* copy number was determined by the sum of *AMY2A, AMY2B*, and *AMY2P*. To assess whether this inter-chromosomal correlation was observed by chance, we randomly sampled 10,000 biallelic loci from the autosomal variants except chromosome 1 where the amylase genes are located, and calculated the correlation between the average copy number of amylase genes and the derived alleles of randomly selected variants. The empirical P-value was calculated based on the distribution of correlation with the randomly sampled variants.

At the transcriptomic level, we analyzed the correlation between amylase and trypsinogen genes with the normalized expression data in the pancreas from GTEx (v8), which contains 22,616 genes from 305 samples. We further performed the weighted correlation network analysis (WGCNA v1.71) ^19^. In the automatic network construction, we set the soft-thresholding power=9. Amylase and trypsinogen genes were clustered in the same module, from which we performed gene ontology analysis for all the genes with Metascape ^63^. For visualization, we filtered out the connection with weight <0.1 and plot the network with Cytoscape (v3.9.1) ^64^.

### Gene expression analysis

We focused on the expression of *PRSS1* and *PRSS2* from the GTEx project (V8) to analyze the impact of the haplotype structure ^20^. For the effect size analysis, we compared rs2855983 (the SNV with complete LD of the copy number structure in Eurasian populations) with other *PRSS2* eQTL-SNVs that were reported in GTEx. In the ASE analysis of the copy number structure, we focused on the samples of European ancestry (n=243) as the composition of the long and the short form haplotypes is almost solely H2 and H1S, respectively. By intersecting the copy number structure surrogated SNVs (R^2^>0.9) with those ASE-SNVs reported in *PRSS2* from the GTEx project, rs2075544 (R^2^=0.97) was selected with reference and alternative allele (GRCh38 primary scaffold) as a surrogate of 3-copy and 5-copy haplotype, respectively. To investigate the haplotype effect on the expression ratio of *PRSS1*/*PRSS2*, we applied the linear regression model to analyze the correlation on two levels: one is between the haplotype configurations and the *PRSS1*/*PRSS2* ratio, and the other is between the ASE effect (the relative expression level between the 3-copy and 5-copy haplotype) and the *PRSS1*/*PRSS2* ratio in the heterozygous 3-copy/5-copy carriers. To compare the correlation between *PRSS1* and *PRSS2* in the three haplotype configurations, we first matched the sample size (n=38) in each haplotype configuration group and randomly sampled 100 times, then chose the sampling group with the mode of correlation coefficient and applied the Jackknife method to compare the coefficient among sampling groups for the three haplotype configurations (Fig. 4a). For expression coefficient between *PRSS1* and *PRSS2* in multiple tissues, we calculated the Pearson’s coefficient for all the available samples in each tissue (Fig. 4c).

### 3D-genome analysis

We investigated the topological associated domain (TAD) of both 3-copy and 5-copy haplotype structures by analyzing the Hi-C data of a homologous 3-copy sample HG00514 and a homologous 5-copy carrier GM12878, of which sequencing reads were available from previous studies ^56,65,66^. The sequencing reads of HG00514 were merged and aligned to CHM13, a high-quality *de novo* assembly of 3-copy haplotype ^67^, while the Hi-C data from GM12878 were self-aligned with the collapsed diploid assembly. We employed HiC-pro (v2.10) ^68^ for sequencing read processing and the contact matrix generation. We used HiCexplorer (v3.7.2) ^69^ for TAD visualization.

### Epigenetic analysis

To search for the potential enhancers at the *PRSS1-PRSS2* locus, we analyzed the histone modification signals with the ChiP-seq data from the ENCODE project ^70^, and aligned the reads to the *de novo* assembly of NA12878 with Bowtie2 (v2.3.3.1) ^71^. We detected the peaks of histone modification H3K4me3, H3K4me1, and H3K27ac with MACS2 (2.1.1) ^72^. The haplotype configuration of these samples was determined by manually checking the read alignment at the haplotype-structure tagged SNVs with Integrative Genomics Viewer (IGV v2.12.3) ^73^.

The H3K27ac peaks were further aligned to the known enhancers of *PRSS1* and *PRSS2* from the Genehancer database ^74^ by lastz. Since the trypsinogen gene copies resulted in multiple alignments due to high sequence similarity, we assigned each H3K27ac peak with the highest similarity in the alignments by S-score, which was defined as F1-score by considering both the precision and recall rate:

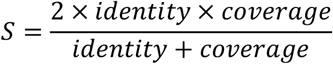

The multiple sequence alignments were visualized by Miropeats, and the alignments with the highest similarity were highlighted in colors.

Given that these H3K27ac peaks are located in the promoter region of five trypsinogen genes, we employed HOMER (v4.11) ^75^ to search for the motifs of a transcription factor that were present in *PRSS3P2, TRY7*, and *PRSS2*, but absent in *PRSS1* and *PRSS3P1*.

### scATAC-seq and scRNA-seq analysis

To explore the regulatory effect of the haplotype structure, we analyzed the scATAC-seq and scRNA-seq data from the same cells in the pancreas and other tissues. The sequencing reads of paired scATAC-seq and scRNA-seq data, which were generated by the 10X Genomics Multiome platform and available for the homologous long-form samples and heterozygous samples, were downloaded from ENCODE project. We aligned the sequencing reads to NA12878 assembly and generated the fragments file and the expression matrix with Cellranger-arc. For the homologous short-form samples, of which only the scATAC-seq data were available, we downloaded the fragments file and converted the GRCh38 coordinates to NA12878 coordinates.

Based on the processed data, we employed ArchR (v1.0.1) ^76^ to perform an integrative single-cell open chromatin accessibility analysis. For the paired RNA-seq and ATAC-seq analysis, we filtered cells that were not enriched in the transcription start site (TSSenrichment ≤ 6) or with fragments less than 2500, we performed Latent Semantic Indexing for dimensionality reduction and clustering. We generated the pseudo-bulk replicates with default settings and called peaks with tile matrix. The ‘peak-to-gene’ links were determined by the correlation between chromatin accessibility and gene expression. The scATAC-seq-only data were analyzed likewise, except for the filtering with default settings, and the ‘coaccessibility’ instead of the ‘peak-to-gene’ link was determined by the correlation of the peaks.

### Mendelian randomization and colocalization analysis

To investigate the causal relationship between the elevated *PRSS2* gene expression (*PRSS1*/*PRSS2* ratio) and the pancreatitis risk, we applied the gene expression data from GTEx and the summary statistics from a previous GWAS study of the European population (n=7,999) ^21^ to perform Mendelian Randomization analysis by using SMR (v1.0.3) ^77^. In addition, an inverse-variance weighted method ^78^ was also used in the Mendelian Randomization analysis with haplotype structure as an instrumental variable, the *PRSS1*/*PRSS2* ratio as exposure, and the disease state of pancreatitis as an outcome.

To identify the causal variant in the gene expression and the GWAS association, we further employed the Bayes Factor colocalization analysis with R package coloc (v5.1.0) ^79^. This analysis can also test whether two associations shared a single common causal variant. The GWAS summary statistics also included the previously reported SNVs that were associated with pancreatitis, such as rs10273639 ^80^ and rs4726576 ^13^. These SNVs were analyzed together with the haplotype structure-tagged SNV rs2855983 to determine the causal variant.

### Pancreas disease association analysis

Most of the previous studies used rs10273639 as a marker to assess the genetic association of pancreatitis at the *PRSS1-PRSS2* locus ^14^, while this SNV is not in complete LD with the copy structure. We used the SNVs rs2855983 and rs13228878 that were in complete LD with the haplotype structure in most of the Eurasian populations to estimate the odds ratio of pancreatitis for the haplotype structure. A total of three studies containing either of the SNVs were included ^21,81,82^. The meta-analysis was performed by R package meta (v4.18) ^83^.

To investigate the susceptibility of the haplotype structure to pancreatitis cancer, we calculated the correlation between the age-standardized pancreas cancer incidents of different ethnic groups from the Surveillance, Epidemiology, and End Results (SEER) Program in the United States and the 5-copy haplotype frequency of the matched ancestry in the 1,000 Genomes Project. The long-form haplotype frequency of Gujarati Indians in Houston (GIH), Han Chinese in Beijing (CHB), Yoruba in Ibadan (YRI), Mexican Ancestry in Los Angeles (MXL), and Utah residents with European ancestry (CEU) from 1,000 Genomes Project were used as surrogates of Indian, Asian, Black, Hispanic and White, respectively.

## Data Availability

All data are publicly available online, see 'Data Availability' section in the manuscript for accession numbers.

## Data availability

All data are publicly available and were download as below: The great ape assemblies from National Center for Biotechnology Information (NCBI) with BioProject number: PRJEB10880 and PRJNA369439; The assemblies of other primates (see Supplementary Table 1 for details) from Ensembl (https://www.ensembl.org/); The short read sequencing data of great ape genomes (https://www.ncbi.nlm.nih.gov/bioproject/?term=PRJNA189439), Neanderthal/Denisovan genomes (http://cdna.eva.mpg.de/neandertal/), modern human samples (see Supplementary Table 2 for details) from 1,000 Genomes Project (https://www.internationalgenome.org/data-portal/data-collection/30x-grch38), and Human Genome Diversity Project (https://www.internationalgenome.org/data-portal/datacollection/hgdp); *De novo* assembly of CHM13 and NA12878 from NCBI GenBank with accession number GCA_009914755 and GCA_002077035.3, respectively; The HiC data from NCBI with accession numbers: PRJNA268125 for NA12878 and PRJNA300843, PRJNA528584, PRJEB39684 for HG00514; The multi-tissue RNA data from GTEx project (release v8; https://gtexportal.org/home/); The sequencing reads of Chip-Seq data from ENCODE project: ENCSR944XDZ, ENCSR761LTB, ENCSR211RGU, ENCSR692WNL, ENCSR857MYG, ENCSR659RJP, ENCSR258LWZ, ENCSR959TTM, ENCSR005DUS; The sequencing reads of 10X Genomics single-cell ATAC and RNA-seq data from ENCODE project ENCSR868CRK, ENCSR229VVY, ENCSR979ZOG, ENCSR281NBH, ENCSR472RRP, ENCSR188TJX. Pancreas cancer incidence: https://seer.cancer.gov/.

## Acknowledgements

This study was supported by the National Natural Science Foundation of China (NSFC) grant (32030020, 31871256, and 31961130380), the Strategic Priority Research Program (XDPB17, XDB38000000) of the Chinese Academy of Sciences (CAS), the UK Royal Society-Newton Advanced Fellowship (NAF\R1\191094), and the Shanghai Municipal Science and Technology Major Project (2017SHZDZX01). The funders had no role in the study design, data collection, analysis, decision to publish, or preparation of the manuscript.

## Author contributions

H.L. and S.X. conceived the study. S.X. supervised the project. H.L. and Y.W. designed the analysis. H.L. and Y.W. performed an evolution analysis. H.L., Y.W., B.X., and R.Z. performed population genetic analysis. H.L., B.X., and X.B. performed multi-omic analysis. H.L., Y.W., B.X., and Y.G. performed data processing. Y.W. and B.X. performed genetic variant calling. H.L. drafted the manuscript. S.X. revised the manuscript.

## Competing interests

None declared.

## Notes

### Competing Interest Statement

The authors have declared no competing interest.

### Author Declarations

The study used ONLY openly available human data that were originally located at NCBI, ENCODE project, 1000 Genomes Project, Human Genome Diversity Project, SEER and GTEx project.

