## Supplementary Information for "Structural evolution of trypsinogen gene redundancy confers risk for pancreas diseases"

**Supplementary Notes**
**Supplementary Fig. 1-22**
**Supplementary Table Legends**

### Supplementary Notes

**Evolution of trypsinogen genes in primates**

To gain an evolutionary insight of human gene duplications at the *PRSS1-PRSS2* locus, we investigated the homologous sequences in primates (Supplementary Fig. 1 and 2). Strepsirrhini, as the primate with the largest genetic divergence to human, has only partial alignments with human trypsinogen gene (identity, 72-76%; coverage, 35-46%). The whole copy alignment can be found in the new world monkeys (identity, ~83%; coverage: 93%), which indicates a single copy origin in primate lineage. The expansion of trypsinogen gene duplication started after the split of the new world monkeys. All species in Catarrhini harbor the duplicated trypsinogen genes, among which the old world monkeys contain at least 9 copies while the apes commonly have 6 copies. To further explore whether the duplications started in the common ancestry of the old monkeys and the apes or occurred independently in the two lineages, we performed Bayesian phylogenetic analysis for the duplication copies. The phylogenetic tree using new world monkeys as an outgroup exhibits a topology of two major branches with one in old the world monkeys and the other in the apes, which suggests that duplications independently emerged in the two lineages (Supplementary Fig. 3b).

In the old world monkeys lineage, the age of the duplication coincided with the its divergence time from the apes. Furthermore, the time to the most recent common ancestor of different copies greatly overlapped, indicating a rapid expansion event. Based on the phylogenetic tree, the trypsinogen genes of the present-day old world monkeys might derive from three ancestral copies since the first expansion around 33 Mya. The second expansion might occur during 27-29 Mya, which evolved to current haplotype structure of 9 to 12 copies (Supplementary Fig. 3b). Although the total copy number varies, the structure of the last 8 copies retains the same across different species.

In the apes lineage, the duplication took place during 24-30 Mya, which also evolved to a 3-copy ancestor around 29 Mya and finally formed a 6-copy structure ~25 Mya (Supplementary Fig. 3b). The major haplotype of present-day great apes retains this structure except gibbon and human. Gibbon has three trypsinogen genes at *PRSS1-PRSS2* locus, and this structure is more likely formed by deletion from the 6-copy structure rather than an ancestral duplication form after the first expansion event (Fig. 1b). This inference was made based on the identity of intergenic sequence at the trypsinogen copies, in which the sequence identity between the orthologous copies of gibbon and other non-human great apes is significantly higher than that between the corresponding duplicated paralogous copies ($P<0.001$, paired Wilcoxon text; Supplementary Fig. 8). If gibbon retains the ancestral form as a 3-copy form of the second duplication expansion since its early divergence from great apes, we would expect that the two duplicated copies of non-human great ape have similar sequence identity to the gibbon's ancestral copy; In fact, we observed significant difference between the two duplicated copies, which supported that the ancestral form of gibbon was also 6-copy and it experienced additional deletion event to form a 3-copy structure. Interestingly, the copies that were lost in the Gibbon lineage all have larger dN/dS ratio than the remaining copies, and in humans these copies became pseudogenized (Fig. 1a and c). We further explored the gene duplication polymorphism in the great apes by aligning short sequencing reads from a previous study [1]. In addition to the 6-copy haplotype, we also observed five chimpanzee samples carried one copy deletion (Supplementary Fig. 6a), i.e., 5-copy haplotype. However, these 5-copy haplotypes are different from ancestral human haplotype, as the deletion occurs at the 3rd copy of the 6-copy structure. Notably, in contrast to high copy number in the old world monkeys (≥9), we observed only deletion polymorphism in the great ape lineage, i.e., no further duplications (copy number >6) were found in either non-human apes or human samples at the *PRSS1*-*PRSS2* locus (Supplementary Fig. 6a). Considering that the structure of this locus is likely to induce non-allelic homologous recombination and that the high copies in human would cause pancreatitis [2], these results suggest that natural selection might act against higher copies of trypsinogen genes in the apes lineage.

To explore the mutations on *PRSS1* and *PRSS2* with signatures of deviating neutral evolution in the apes lineage, we performed Bayes Empirical Bayes method to detect natural selection (Methods). Four sites (3P, 11A, 102K and 148A) showed significant signals (df=2, p<2e-08). The 3P and 11A located on the signal peptide region of trypsin family, while the 102K and 148A located on Alpha-trypsin chain 1 and Alpha-trypsin chain 2, respectively. However, the active sites and metal binding sites are highly conserved across multiple copies of trypsinogen genes in all primates.

**Haplotype evolution of human trypsinogen genes**

To infer the possible ancestral state of human haplotype, we compared the present-day human sequences with the chimpanzee sequence. We counted the nucleotide difference between human and chimpanzee with random sampling 100 sequences in each haplotype group for each duplicated copy, except the chimpanzee *PRSS1* region, which was manually assembled with 1.78kb and 8.9kb chimpanzee sequence from the first and the second duplicated copy, respectively. The nucleotide difference between human and chimpanzee varied from 170 to 330 bases across different duplicated copies. When comparing different haplotypes in each duplicated copy, the nucleotide differs 10 to 30 bases across groups. Notably, different human haplotypes sharing the same motifs have similar distances to the chimpanzee (Supplementary Fig. 12). For example, sequences with H2-specific motif (H2, H3, H5-2, H5-3, H5-R3 and HR3) have smaller divergence than other haplotypes with non-H2 motifs at the 1st copy. Based on parsimony, we reconstructed the ancestral state of human sequence with the haplotype group that showed smallest nucleotide difference to the chimpanzee sequence. This resulted a human ancestral sequence with the motif feature of H2-H1-H2-H2-H5 for copy1 to copy5 (Supplementary Fig. 10c).

To infer the haplotype evolution history, we constructed the phylogeny of long-form haplotypes in the African population. The phylogenetic trees consistently supported that the earliest divergence emerged between H3 and other haplotype groups (Supplementary Fig. 13), largely due to the Yin-Yang haplotype structure at the *PRSS3P1* and *PRSS3P2* regions between H3 and non-H3 groups (Supplementary Fig. 11), which was defined as two major haplotypes with different alleles at every SNV site [3]. We named the H3 haplotype as Yin-lineage, which includes the alternative contig in GRCh38 (KI270803v1). All the non-H3 haplotypes were named as Yang-lineage. The divergence time between H3 and non-H3 groups was estimated from 0.71-1 Mya. Haplotypes of archaic hominid all belong to the Yin-lineage, in which both Altai and Vindijia Neanderthals carried the H3-like haplotype and separated from modern human ~600 kya. Despite of the loss of *TRY7* copy, the alleles at the remaining copies of Denisovan sequences are H3-like (Supplementary Fig. 11). In addition, the African-specific 4-copy haplotype also belong to theYin-lineage, with difference that the deletion occurs at the 2nd copy (*PRSS3P1*). Both of the 4-copy structures are likely caused by non-allelic homologous recombination (Supplementary Fig. 6d), and they exclusively emerged in the Yin-lineage, suggesting this sequence structure might be prone to NAHR.

In the Yang-lineage, the H5 super-group has the largest TMRCA, among which H5-R3 has the deepest coalescence (Supplementary Fig. 13), suggesting that it could be an ancestral form of the Yang-haplotypes. Under the ancestral H5 super-group, there are two major sub-lineages, i.e., H1/H4-lineage and the H2-lineage (Supplementary Fig. 13). In the H2-lineage, the major haplotypes are H5-2 and H2, the latter of which is the most common long-form haplotype in the non-African population, and might be generated by the recombination of H5-2 and H3 haplotypes around 83-126 kya. In H1/H4 lineage, H1 was derived from H5-1 and likely involved in the recombination with H5-4, H3, and H5-2, which resulted in H4, HR1 and HR3, respectively. The deletion event that contributes to half of the present-day short-form haplotypes also emerged on H1 haplotype around 74-260 Kya. Although other short-form haplotypes H4S and HR3S share the same deletion of *PRSS3P2* and *TRY7* copy with H1S, they were derived from recombination of H1S with H4 and H2 respectively, rather than independent deletion events, which is supported by these following lines of evidence: i) the breakpoints of these haplotypes are identical; ii) these haplotypes share the similar haplotype pattern at *PRSS3P1* and *PRSS2* copies (Supplementary Fig. 10b); iii) the pairwise sequence difference of H4S is smaller than H1S (Supplementary Fig. 14); iv) the divergence from H4S and HR3S to H1S is smaller than that from H4S and HR3S to any other long-form haplotypes; v) H1S is the major short-form haplotypes in the African population. Collectively, the H1S is the most common ancestor of the short-form haplotypes.

In the non-African populations, the haplotype diversity was dramatically reduced. The major forms are H2, H1S, HR2, H4S and HR3S, which consist ~96% of the sequences. All these haplotypes were African origin except HR2, which is a recombined form of H1S and H2 that occurred 61-79kya. HR2 distributed in European, South Asian and East Asian populations but absent in Africans, which is the only non-African-specific haplotype that might emerge during the early out-of-Africa.

For both *PRSS1* and *PRSS2* genes, the major amino acid sequences of the long-form and short-form haplotype structure show no difference. However, two haplotype structures show different non-synonymous mutation distribution patterns, and very few of the non-synonymous sites are shared by the two haplotypes (5/41). In contrast to the enrichment of non-synonymous mutation at *PRSS2* (16.3%; 282/1728) compared to *PRSS1* (0.6%; 10/1728) in the 5-copy haplotypes, the non-synonymous mutation distributed evenly between *PRSS1* (1.8%, 42/2318) and *PRSS2* (2.2%; 51/2318) in the 3-copy haplotype (Fig. 1d). Such difference could be explained by the older age of the long-form haplotype and the relaxed selection pressure on *PRSS2*. Moreover, the most common non-synonymous mutation sites at the two protein-coding genes located in two different haplotypes, with *PRSS1* missense variants on the 3-copy haplotype and *PRSS2* missense variants on the 5-copy haplotype. Notably, a previously reported pancreatitis-protective mutation G191R [4] is exclusively located on H2 and segregated in the Eurasian populations (Fig. 1d).

### Supplementary Figures

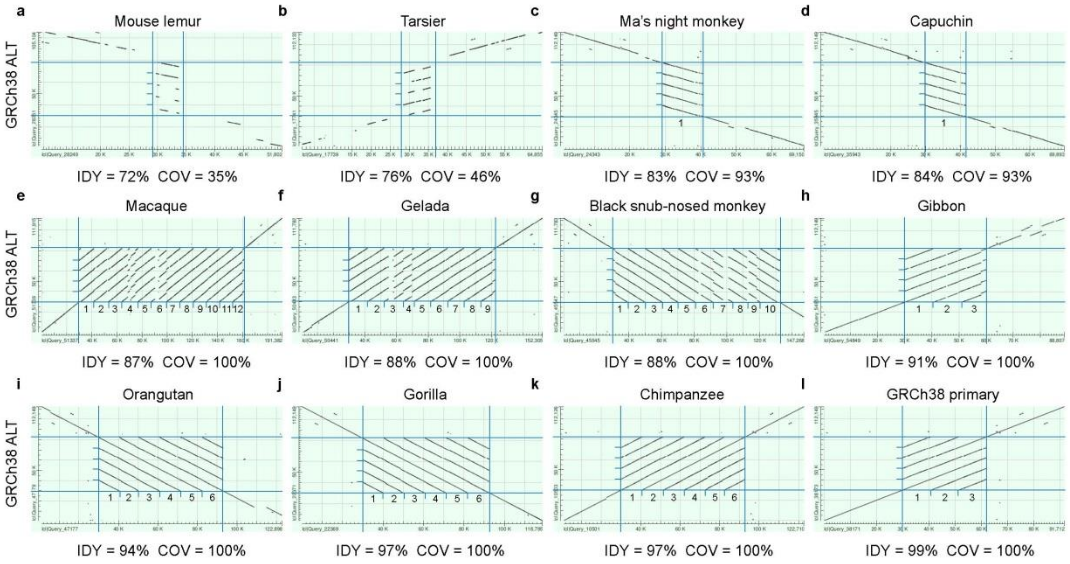


**Supplementary Fig. 1. Sequence alignment of trypsinogen genes between human 5-copy sequence and other primate sequence.**

The vertical axis represents the 5-copy trypsinogen gene structure in human reference genome GRCh38 (chr7_KI270803v1_alt:749409-801557). The horizontal axis represents the homologous primate sequence. Blue lines indicate the alignments between species and the numbers indicate tandem duplicated copies in the primates. IDY, identity; COV, coverage.

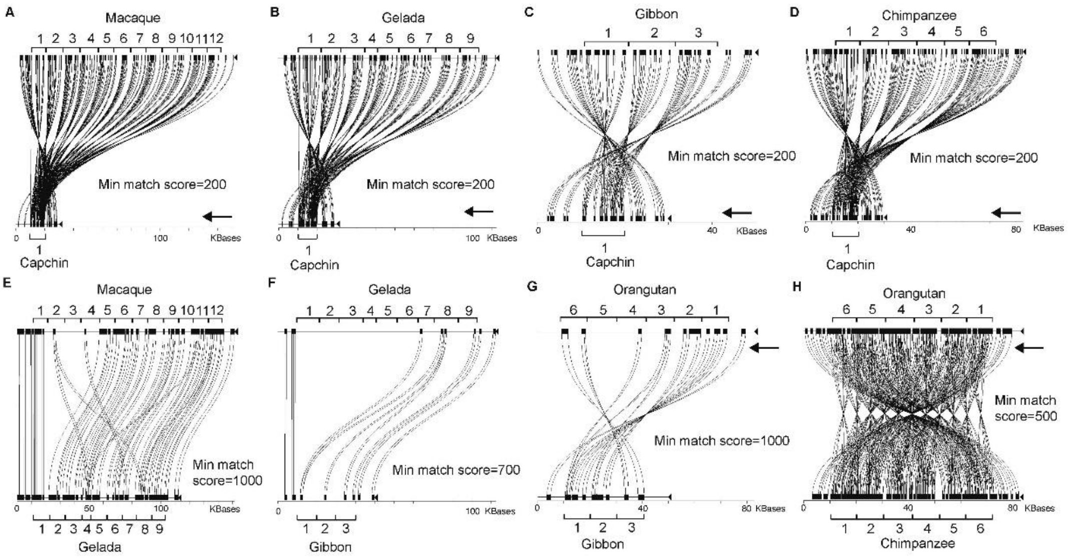


**Supplementary Fig. 2. Sequence alignment of trypsinogen genes between non-human primates.**

Miropeats plot of sequence alignments. The lines and curves show the aligned sequences under different min match score settings. The arrow indicates reverse complement sequence. The numbers indicate tandem duplicated copies in the primates

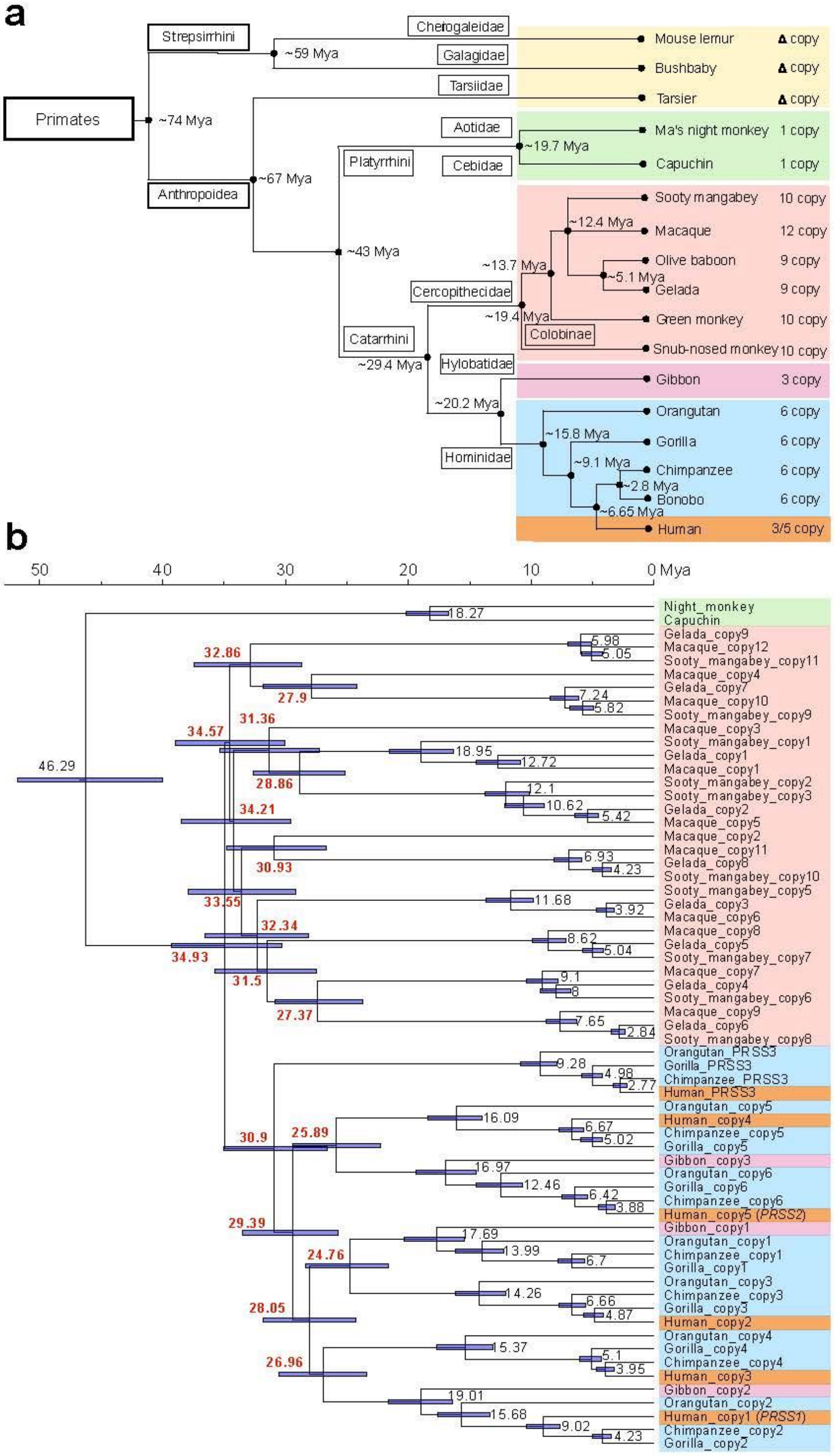


**Supplementary Fig. 3 Phylogenetic tree of trypsinogen genes in primates.**

**a.** Species tree of primates with divergence time obtained from TimeTree. The trypsinogen gene copy number at *PRSS1-PRSS2* locus are shown next to the species name. Delta signal represents partial alignment. **b.** Bayesian phylogenetic tree of different trypsinogen gene copy (including both genic and intergenic regions). The duplication time are highlighted in red. See Supplementary Table 1 for positions of each copy.

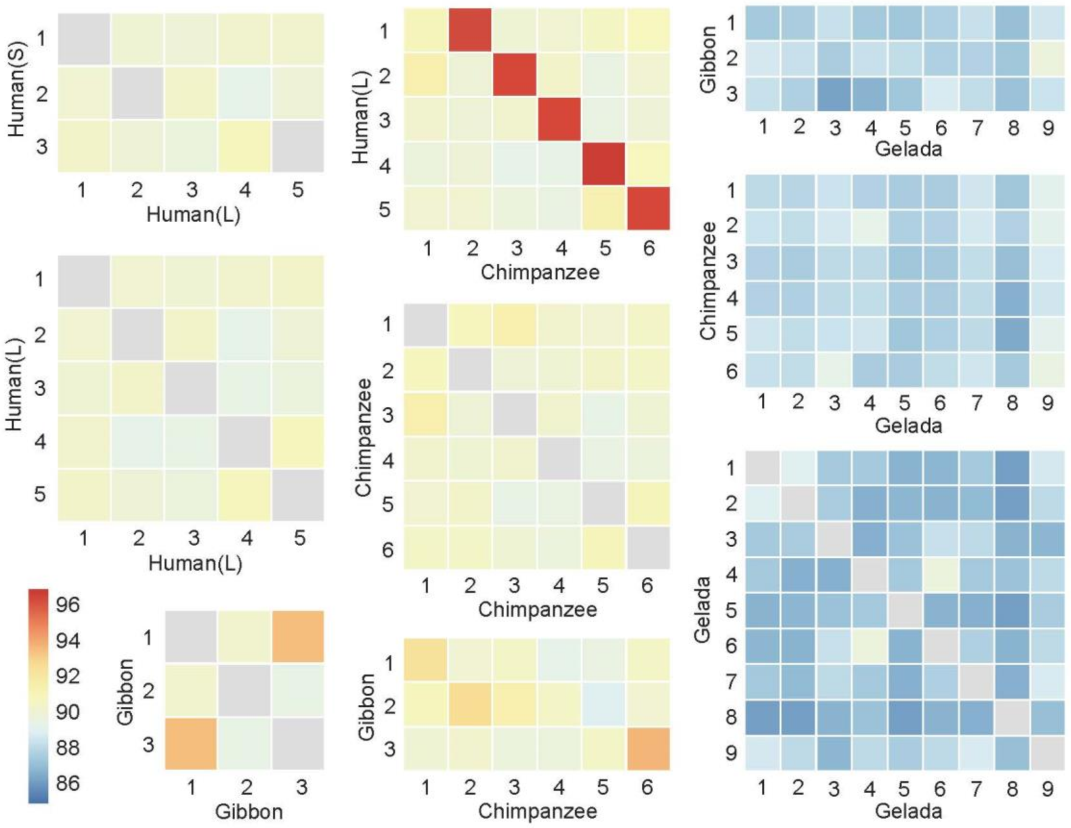


**Supplementary Fig. 4 Sequence identity between different trypsinogen gene copies.**

The heatmap of sequence identity of trypsinogen genes for intra- and inter-species in human, chimpanzee, gibbon and gelada. The intra-species self-alignment in the diagonal element (identity >99%) is marked in gray for display purpose.

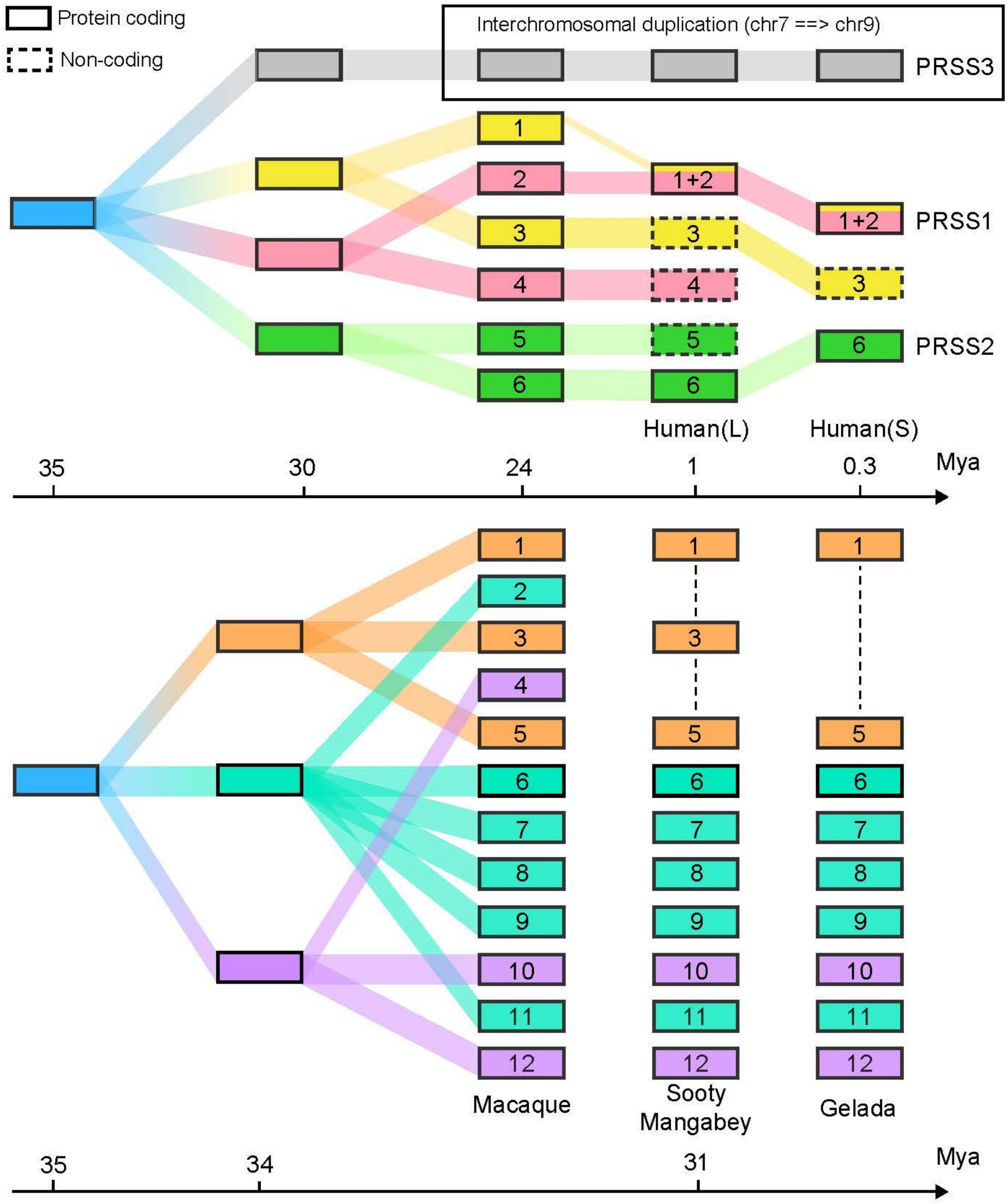


**Supplementary Fig. 5. Diagram of trypsinogen gene duplication evolution in primates.**

This duplication model refers to the 10.6-kb trypsinogen gene copy (including both the protein-coding sequence and the intergenic sequence). Based on the phylogenetic tree (Supplementary Fig. 3), duplication occurred independently in apes (upper panel) and old world monkeys (lower panel). The duplicated copies at the trypsinogen gene cluster (*PRSS1-PRSS2* locus) might have experienced a 3-copy form after the first wave of expansion in both the apes and old world monkeys lineages.

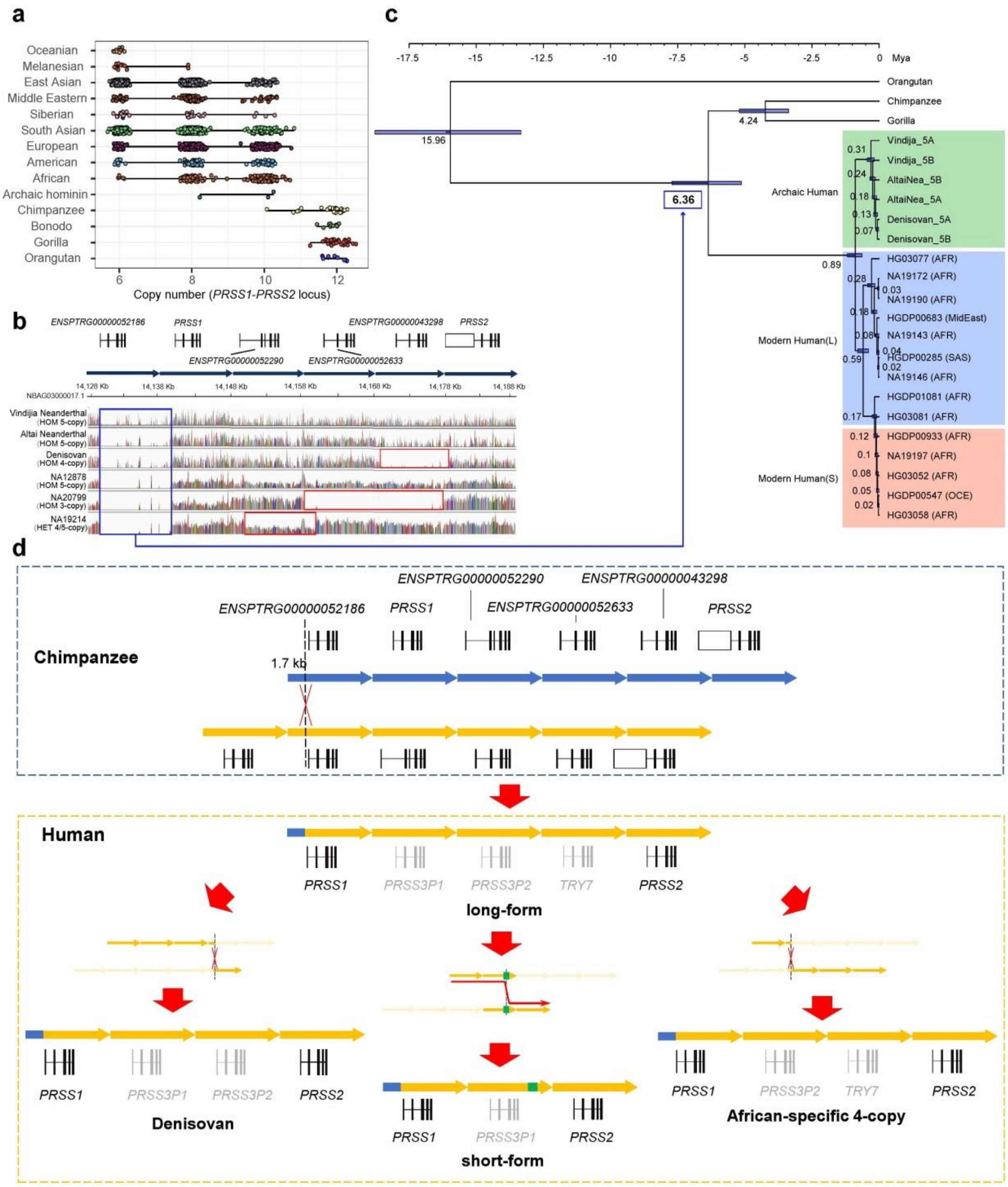

**Supplementary Fig. 6. Human-specific deletion and short-reads alignments of archaic and modern human.**

**a.** Diploid gene copy number of great ape samples at the trypsinogen gene cluster on chromosome 7 (*PRSS1-PRSS2* locus). **b.** Short read alignment of archaic and modern human samples to chimpanzee assembly. NA12878 and NA20799 are the modern human with homologous long-form and short-form haplotype, respectively. NA19214 is an African heterozygous 4/5-copy carrier, which has an African-specific deletion of chimpanzee third copy. The blue frame denotes the human-specific deletion and the red frames denote the polymorphic deletion in the human lineage. **c.** Phylogenetic tree of the human 1$^{st}$ copy (*PRSS1*) and the fused copy of non-human great apes. Time in the blue box indicates the time estimate of fusion event. **d.** Homologous recombination induced trypsinogen gene deletion in human lineage. The blue box shows a non-allelic homologous recombination (NAHR) model that explains the structural difference between chimpanzee and human sequence at the trypsinogen gene cluster. The NAHR occurs at the centromeric 1.7 kb of the first copy and the telomeric 8.9 kb of the second copy of the six-copy sequence in the common ancestor of chimpanzee and human. However, the genic region of human *PRSS1* is not a fusion gene, as the gene body is embedded in the 8.9 kb of the second copy in the common ancestor of chimpanzee and human. The orange box shows how the long-form haplotype evolved into 4-copy haplotype in Denisovan and African population by NAHR and the long-form haplotype evolved into short-form (3-copy) haplotype by homologous recombination [probably mediated by LINE (green)]. The genes in black represent the protein-coding genes and those in gray represent the pseudogenes.

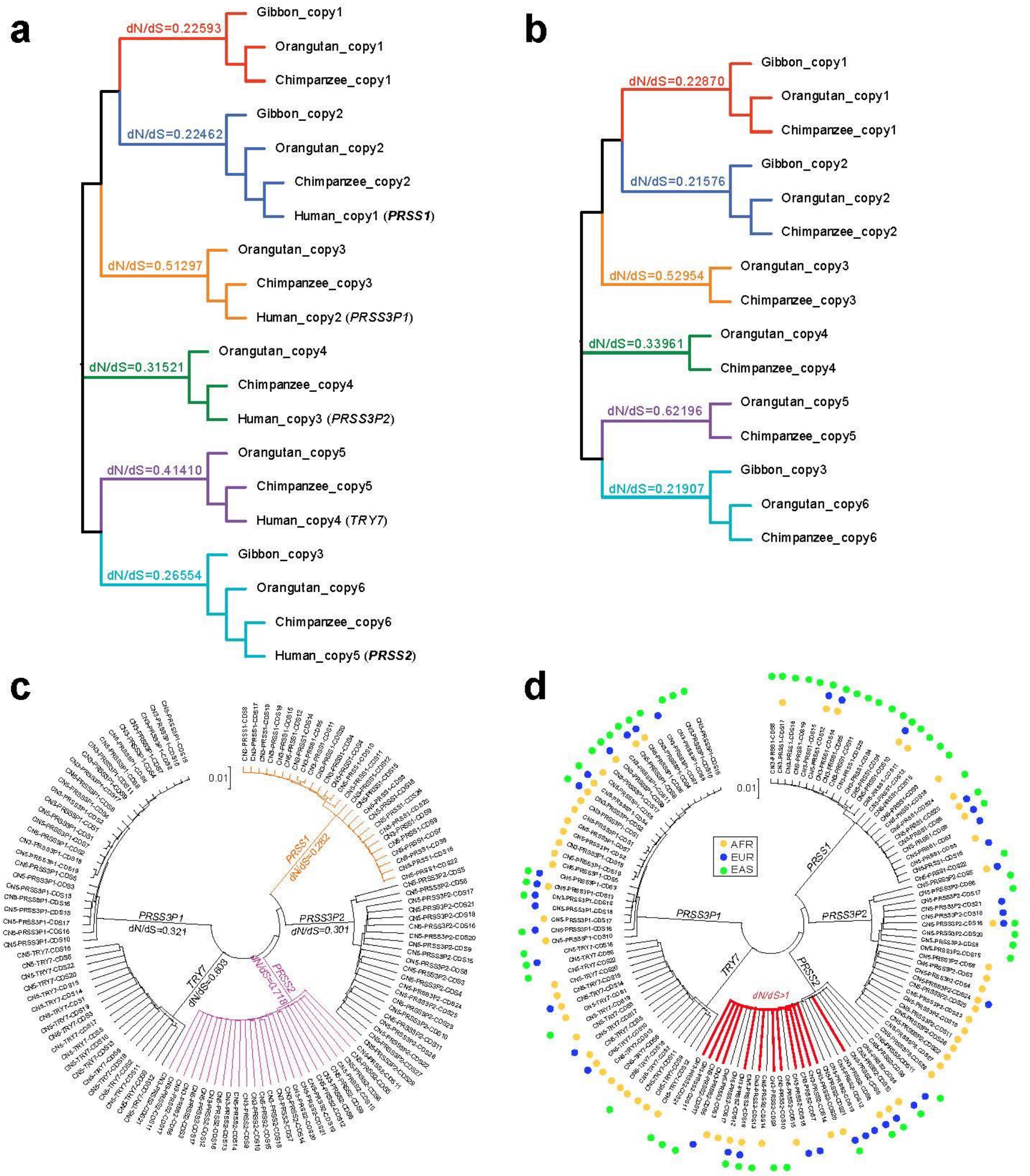


**Supplementary Fig. 7. Natural selection test of trypsinogen coding sequence.** Branch model of dN/dS test for coding sequences of trypsinogen genes in great apes (**a**), non-human great apes (**b**), modern humans (**c** and **d**). The phylogeny in (**a**) and (**b**) is constructed by the entire trypsinogen gene copy (including genic and intergenic regions). The phylogeny in (**c**) and (**d**) is identical, which is constructed by the coding sequences of polymorphic haplotypes in modern human. The foreground branches in (**c**) are the trypsinogen genes such as colored in *PRSS1* and *PRSS2*, while the foreground branches in (**d**) are different combinations at each of the trypsinogen gene with haplotypes from a given population. The outer color circle represents the haplotype that was observed in different populations: orange, African; blue, European; Green, East Asian. For example, the branches highlighted in red at *PRSS2* represent the foreground branch with *PRSS2* coding sequence observed in any one of the African, European and East Asian populations. See Supplementary Table 4 for the ID of coding sequences in (**c**) and (**d**).

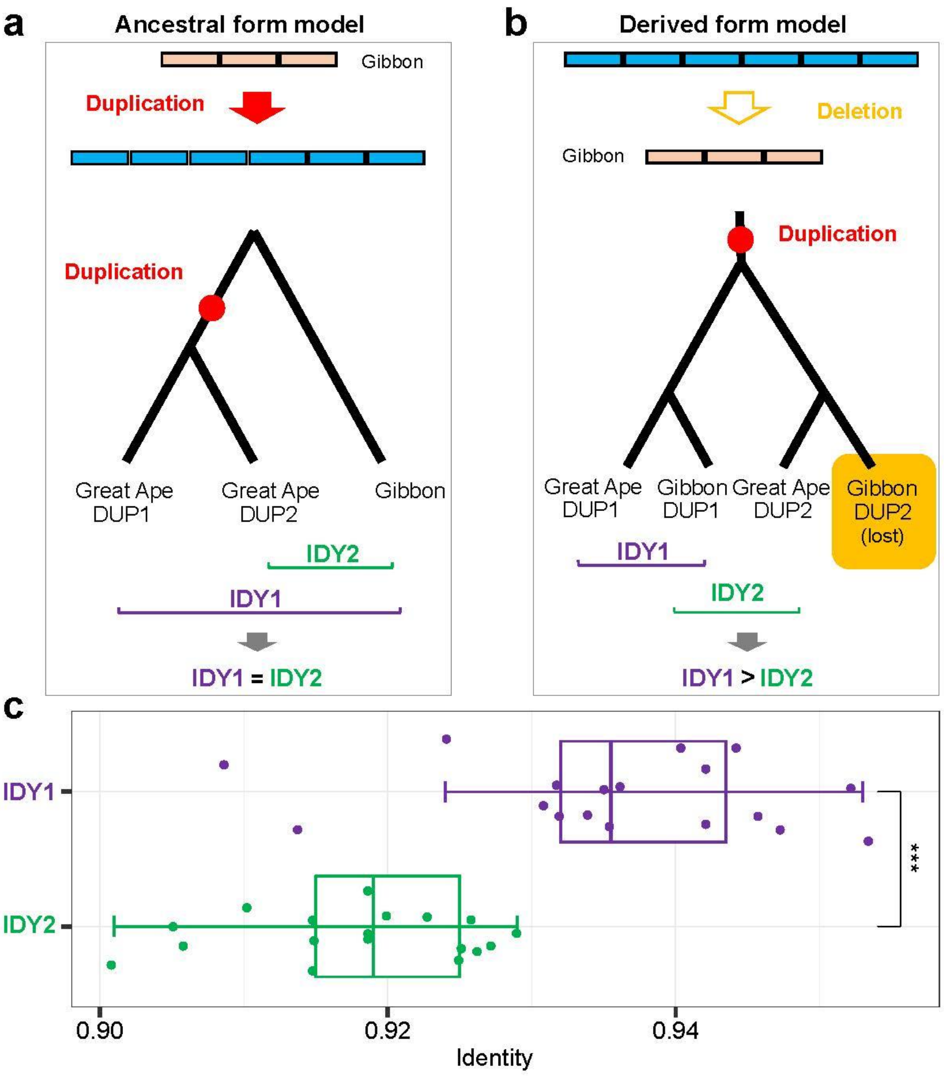


**Supplementary Fig. 8. The evolution model of trypsinogen genes in gibbon.**
**a.** Gibbon retains an ancestral form of 3-copy since the divergence with great apes (Supplementary Fig. 3). In this model, duplication from 3-copy to 6-copy occurs specifically to the great apes, which leads to similar sequence identity between the copy in gibbon and the two duplicates in great apes. **b.** Gibbon had a common ancestral form of 6-copy with great apes and underwent additional deletion. In this model, the asymmetric identity occurs between gibbon and great apes, with the larger sequence identity found in orthologous pairs. IDY, identity. c. Observed identity difference of intergenic sequences at trypsinogen gene copy supports a derived model (**b**).IDY, identity; ***, P<0.001.

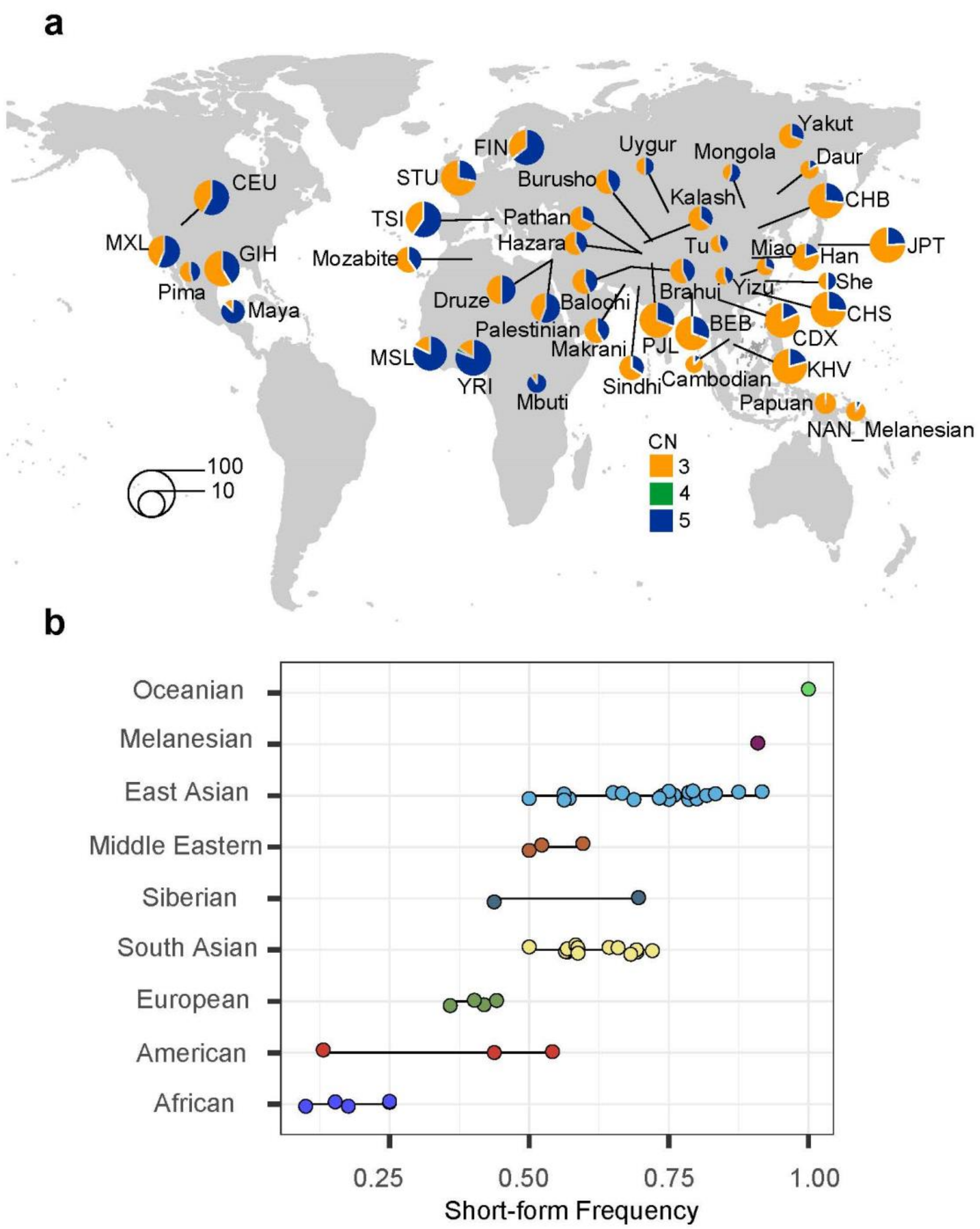


**Supplementary Fig. 9. Frequency of long-form and short-form haplotypes in worldwide populations.**

**a.** Frequency distribution of different copy number structure at *PRSS1-PRSS2* locus in worldwide populations. **b.** Short-form haplotype frequency in super-populations.

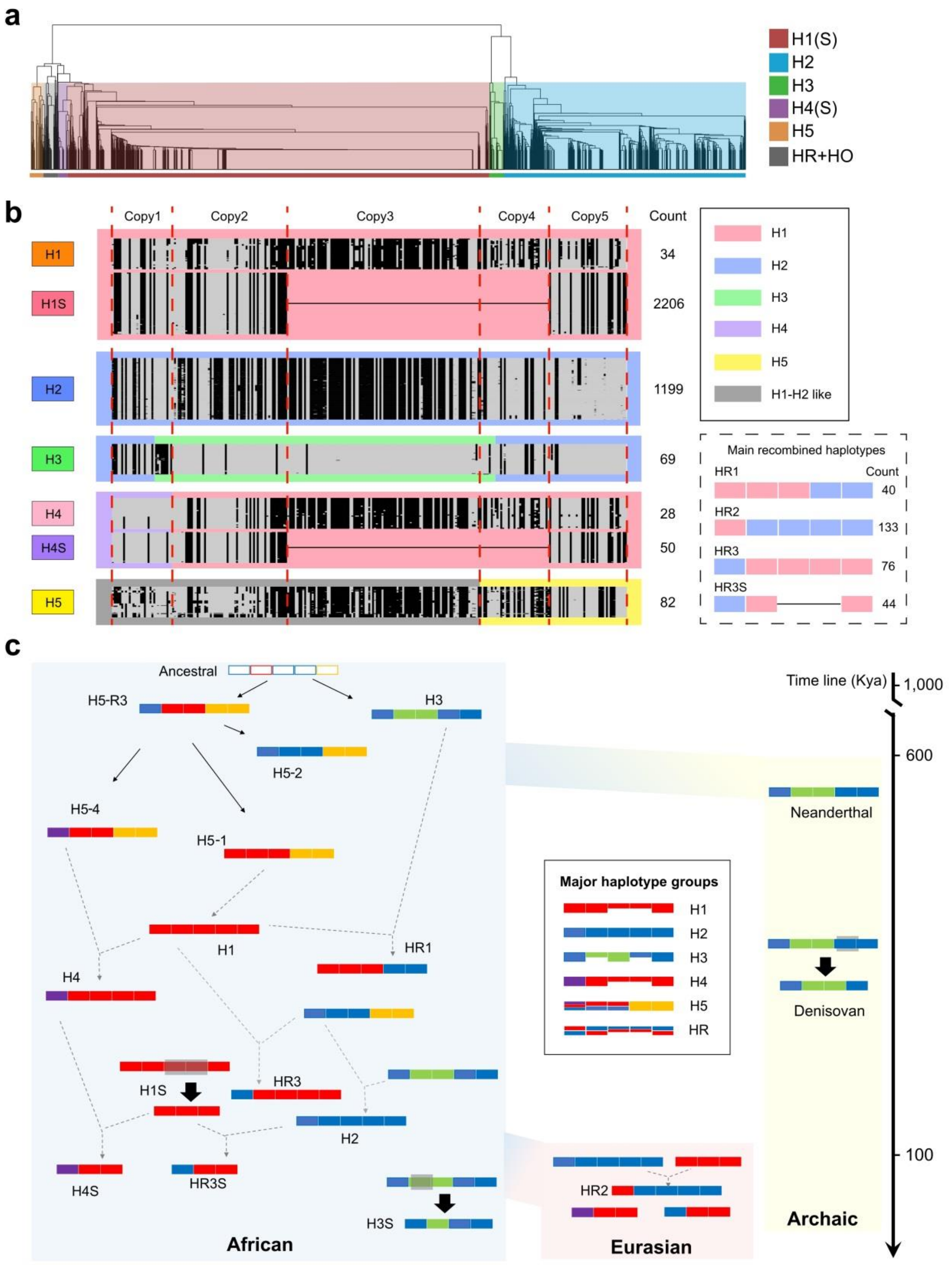


**Supplementary Fig. 10. Haplotype classification and evolution in modern human.**

**a.** UPGMA clustering of haplotypes with variants of MAF>0.01 in 2,027 worldwide samples. **b.** Haplotype pattern in each haplotype group. Each line represents an individual haplotype. In each haplotype box, the reference and alternative allele with respect to GRCh38 alternative contig (chr7_KI270803v1_alt) was indicated by grey and black, respectively. The width of each copy corresponds to the number of called variants. The group-specific sequences were indicated by different colors of the outer frame. **c.** Evolution history inference of human haplotypes (See Supplementary Notes for details).

Solid line indicates the process possibly mediated by mutation, dash line indicates the process mainly mediated by recombination.Shaded grey box indicates deletion event.

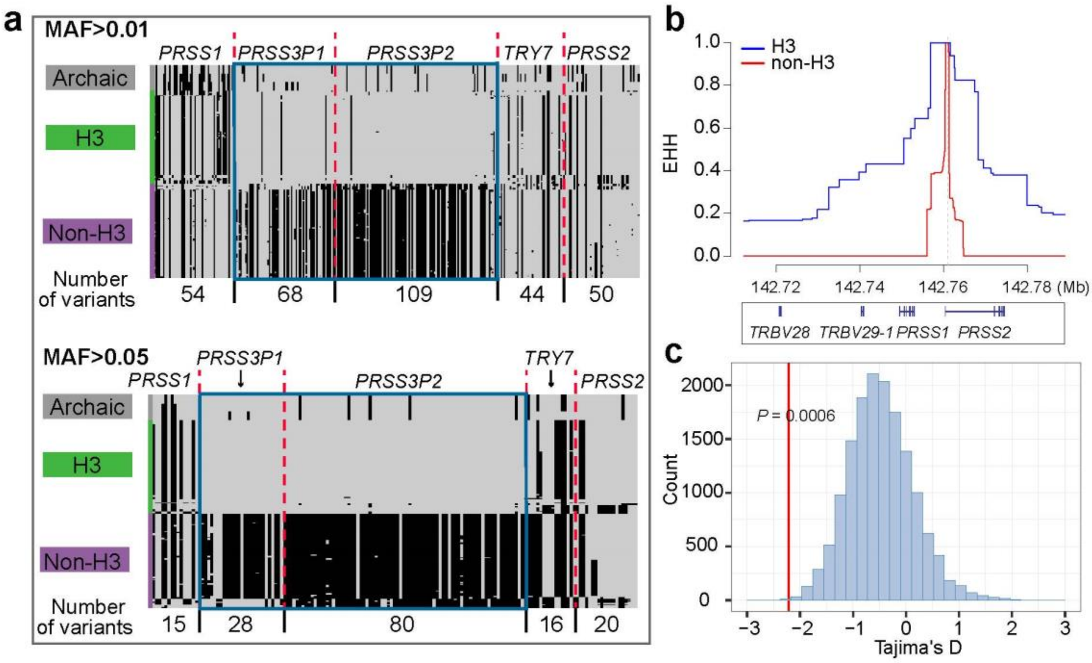


**Supplementary Fig. 11. The Yin-yang haplotype and positive selection on H3 haplotype.**

**a.** Each line represents a haplotype and each column denotes a variant satisfying MAF>0.01 (upper panel) and MAF>0.05 (lower panel). The reference (GRCh38 KI270803v1) and alternative allele of the variant is in gray and black color, respectively. The boundaries of the trypsinogen gene duplicates are separated with red dash line. The blue frame denotes the typical region of Yin-yang pattern that the consecutive variants have totally opposite alleles. The archaic hominids have a H3-like haplotypes. **b.** Extended haplotype homozygosity (EHH) of the H3 and non-H3 haplotypes. **c.** The H3 haplotypes at GRCh38 primary chromosome 7 regions show a significant negative Tajima's D value compared with a background distribution of all haplotypes (including the short-form haplotypes) with 30-kb window.

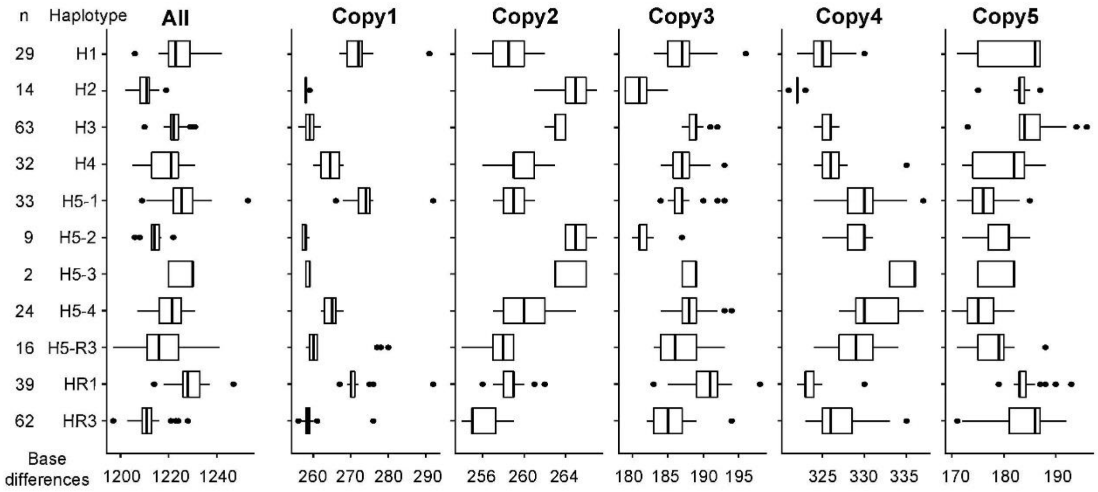


**Supplementary Fig. 12. Divergence between chimpanzee and human in nucleotide at *PRSS1-PRSS2* locus.**

The boxplot shows the nucleotide difference between chimpanzee and human for each trypsinogen gene copy. The number of copies corresponds to the human structure. Copy1 here in chimpanzee was manually fused according to NAHR model (Supplementary Fig. 6).

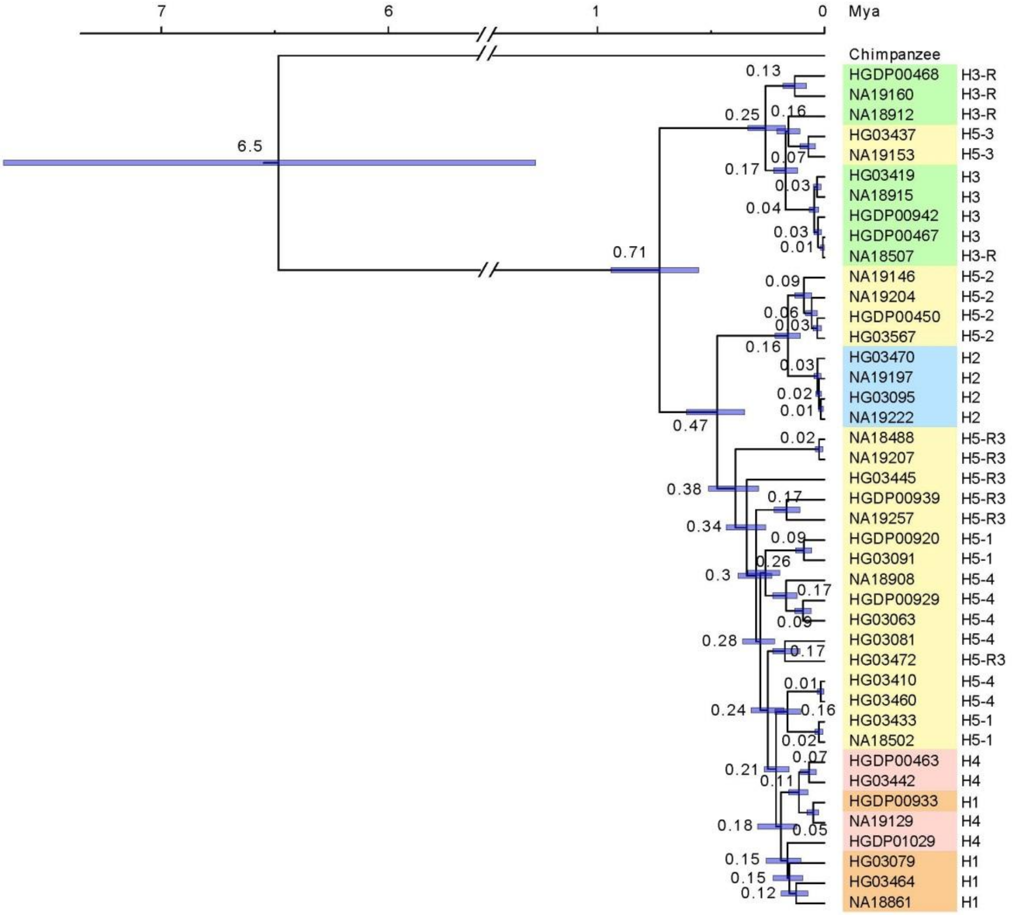


**Supplementary Fig. 13. Phylogenetic tree of long-form haplotypes in African population.**

Four haplotypes from each subgroup were selected to construct the phylogeny except H5-3 which only has two samples. The 52-kb full sequences at *PRSS1-PRSS2* locus were used in the analysis.

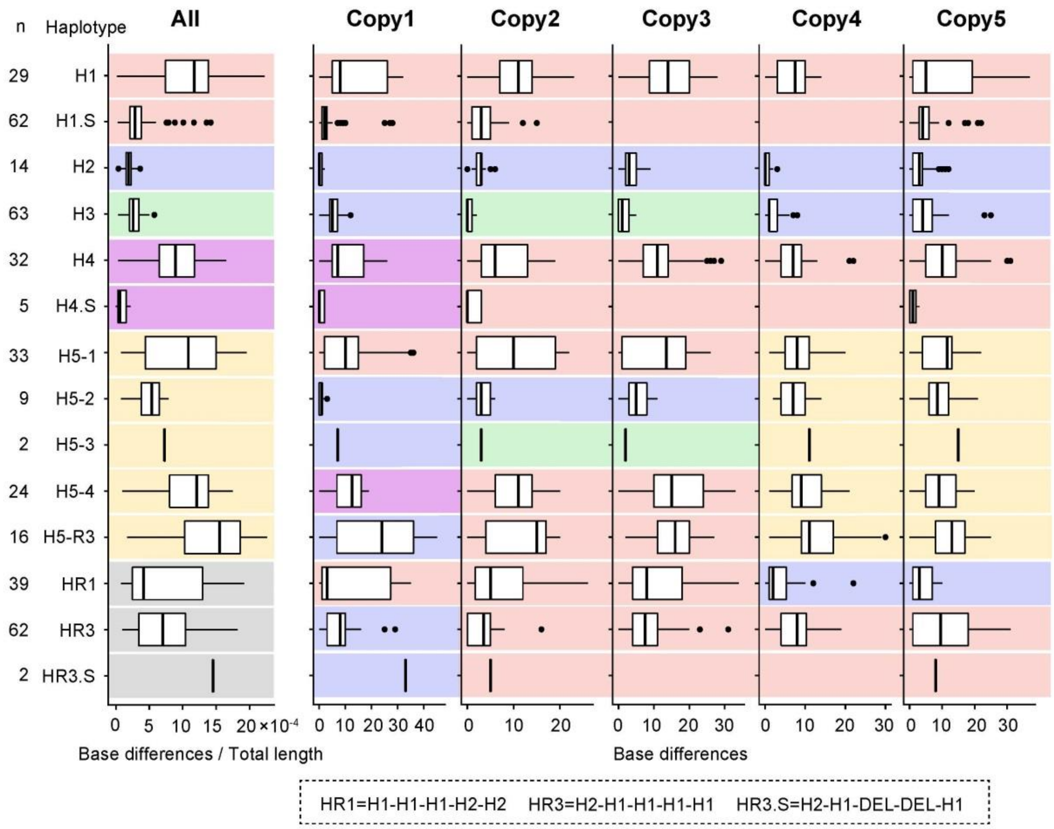


**Supplementary Fig. 14. Pairwise nucleotide difference of each haplotype group in the African population.**

Nucleotide difference for a pair of haplotypes in the same group. Because of the presence of the short-form haplotype, the density of nucleotide difference was calculated when compared all five copies (the "All" panel on the left).

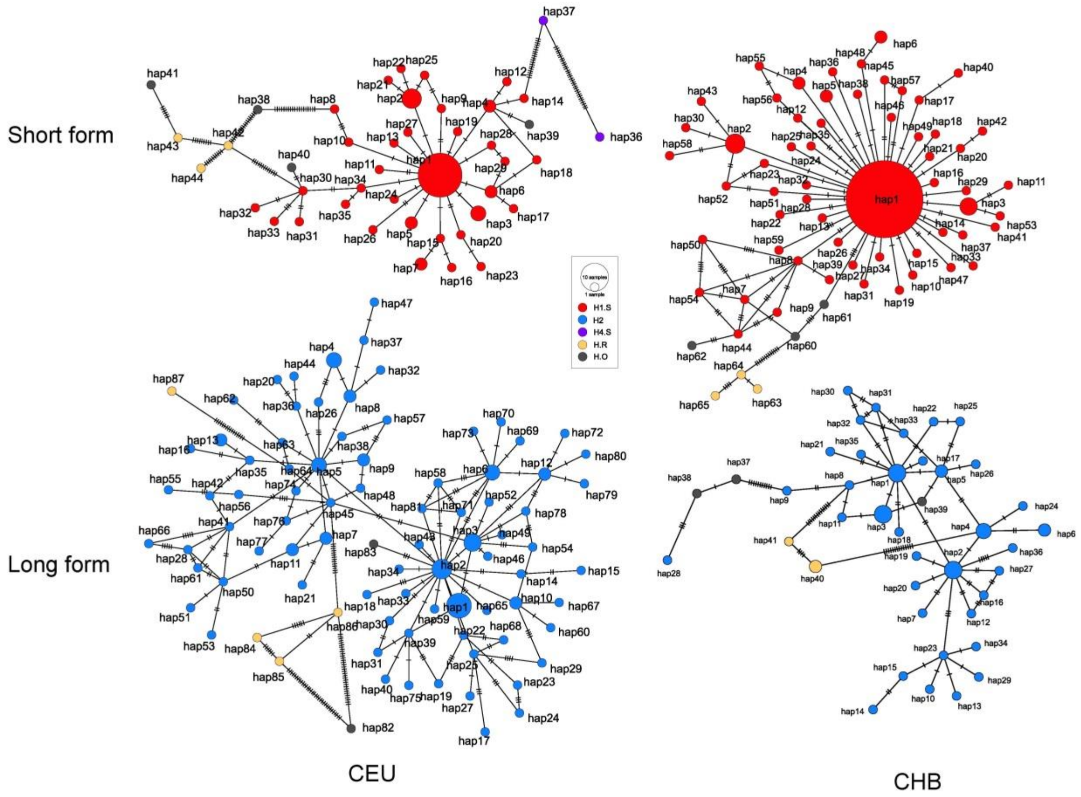


**Supplementary Fig. 15. Haplotype-network in the European and East Asian populations.**

The short-form (upper panel) and long-form (lower-panel) haplotype network in Utah residents with European ancestry (CEU) and Han Chinese in Beijing (CHB). The short-form haplotype network shows a star-like pattern in both CEU and CHB.

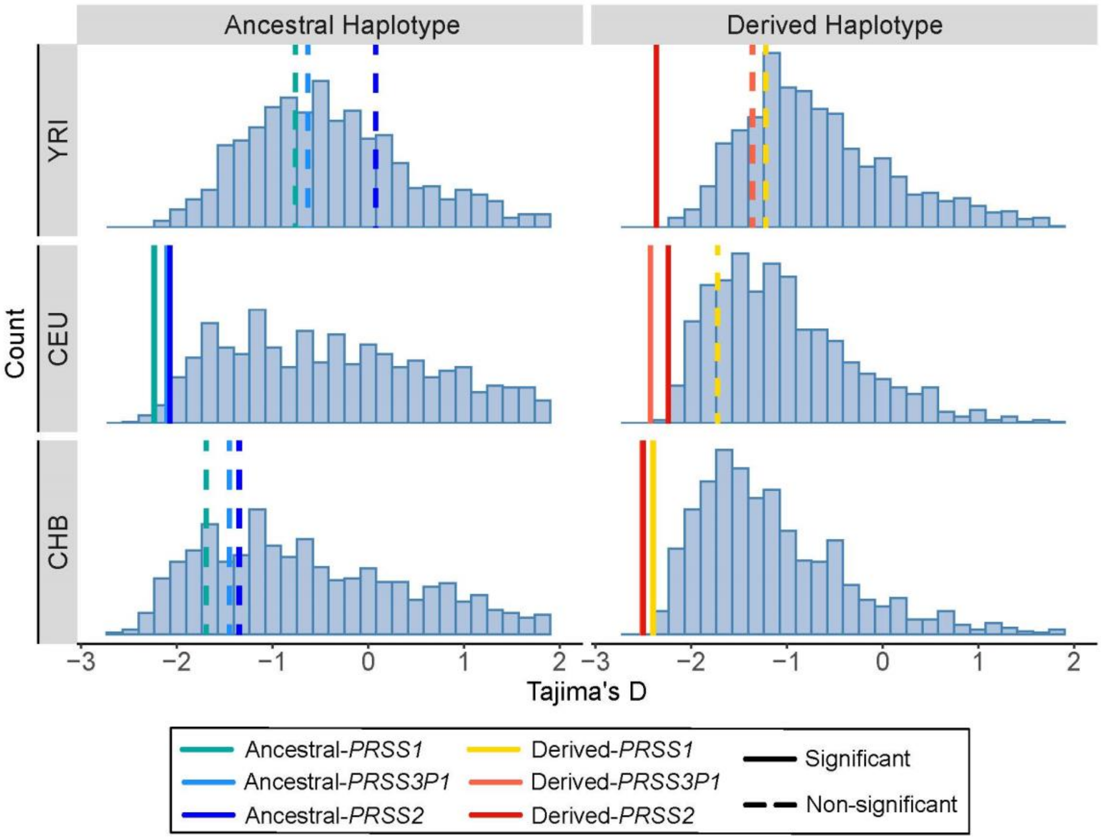


**Supplementary Fig. 16. Tajima's D distribution based on the demographic simulation.**

The histogram shows the distribution of Tajima's D based on the demographic simulation, which follows a three population Out-of-Africa model from ref. [5]. The observed real data were shown in color. Tajima's D was calculated based on 10-kb windows.

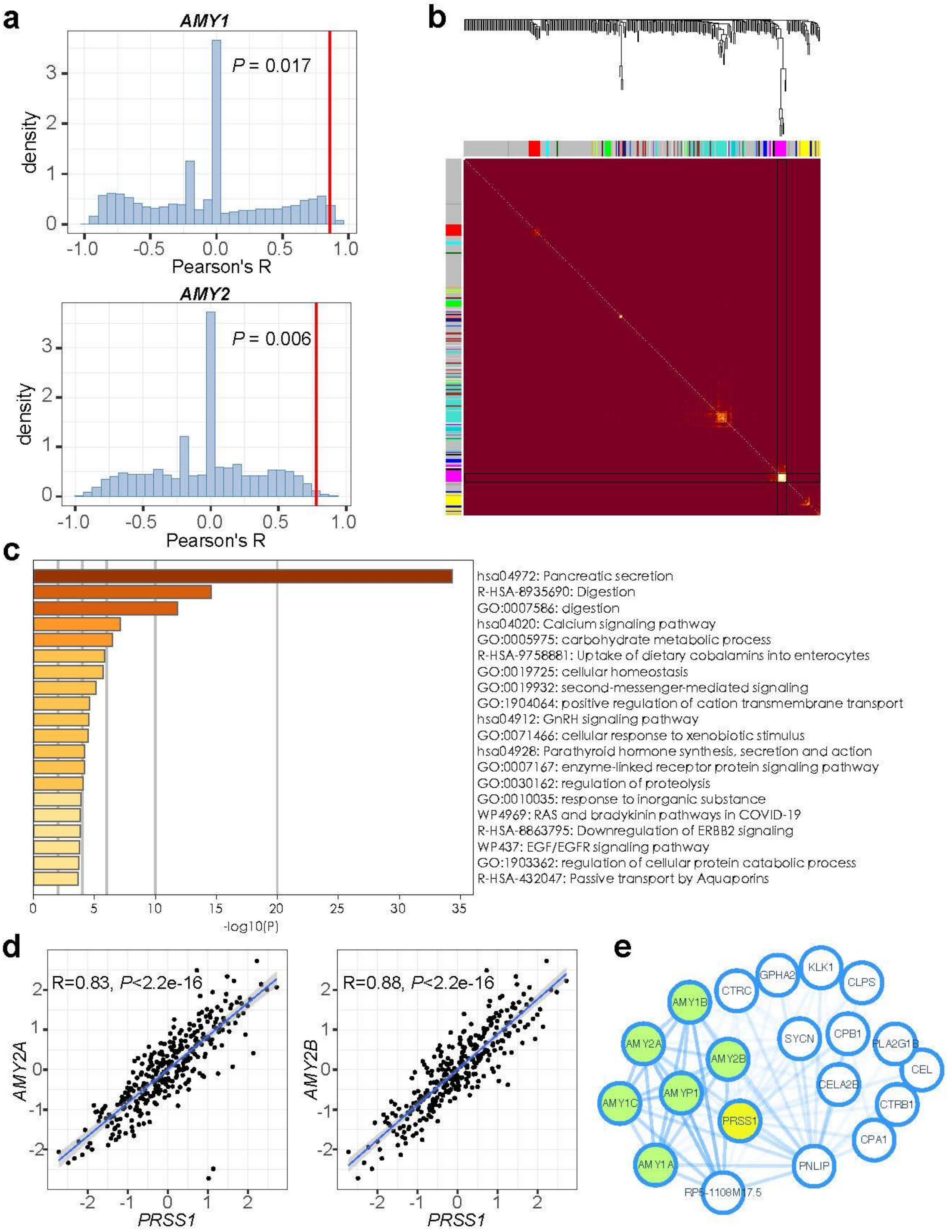


**Supplementary Fig. 17. Genomic and transcriptomic correlation between amylase and trypsinogen genes.**

**a.** The distribution of correlation coefficient between derived allele frequency of randomly selected 10000 variants from chromosome 2 to 22 and amylase gene (*AMY1*, salivary amylase gene, upper panel; *AMY2*, pancreatic amylase gene, lower panel). Red lines denote the observed short-form frequency of trypsinogen gene structure. **b.** Heatmap of amylase and trypsinogen genes together with 300 randomly selected genes in different modules constructed by WGCNA in pancreas tissue. Amylase and trypsinogen genes are all in the module in magenta, which have higher intra-module correlation (yellow in heatmap) compared with genes in other modules (red in heatmap). **c.** Gene ontology analysis of module in magenta shows enrichment in pancreatic secretion. **d.** Pancreatic gene expression between *AMY2* (*AMY2A* and *AMY2B*) and *PRSS1*. **e.** Network plot of module in magenta with genes that have connection weight≥0.1. The amylase and trypsinogen genes are highlighted in green and yellow, respectively. The thickness of the edges represents the connection weight.

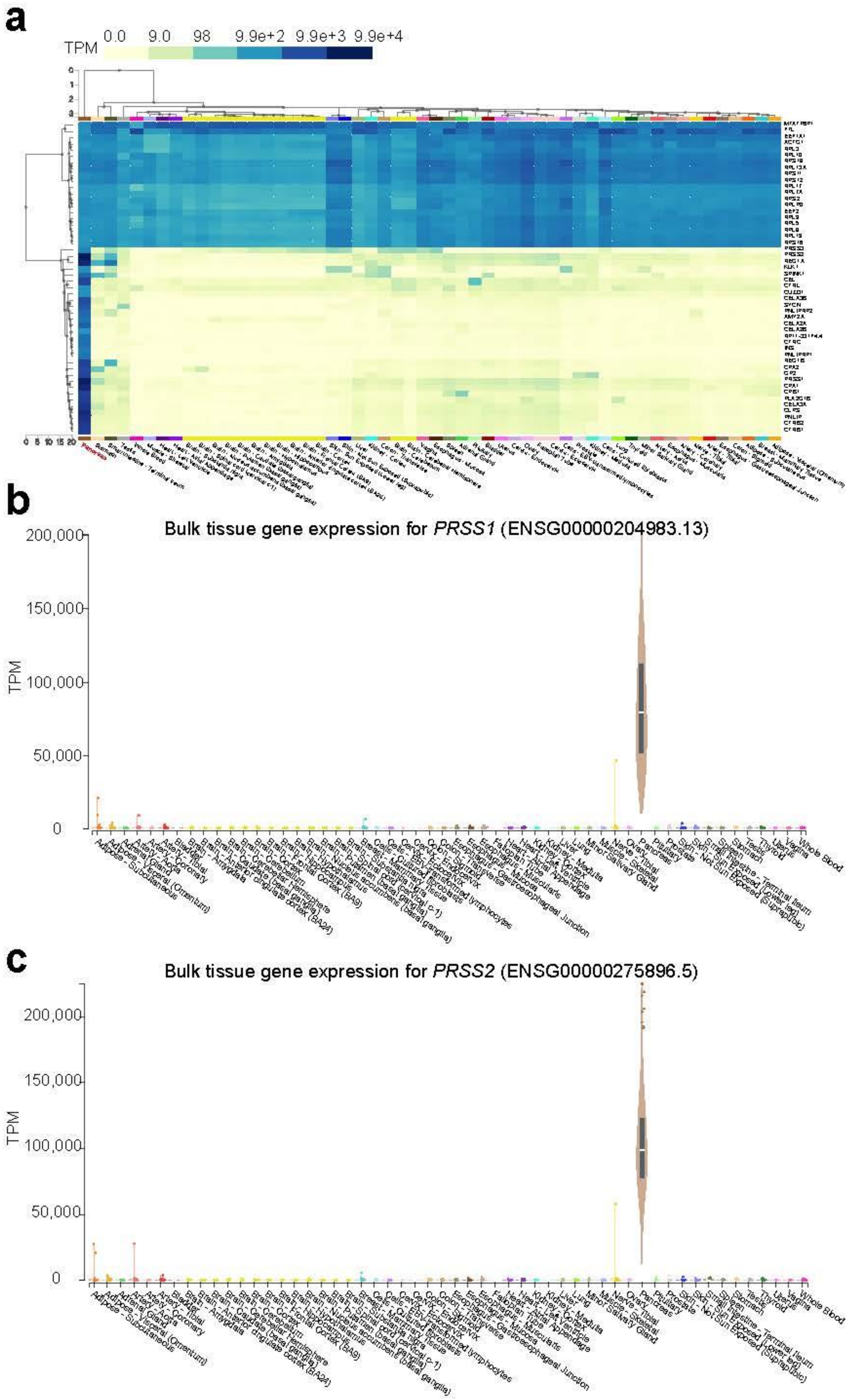


**Supplementary Fig. 18. PRSS1 and PRSS2 expression in multiple tissues.**

**a.** Top 50 expressed genes in pancreas; *PRSS2* and *PRSS1* are the two top expressed genes in pancreas. **b.** *PRSS1* expression in different tissues. **c.** *PRSS2* expression in different tissues. Data from GTEx portal (https://www.gtexportal.org).

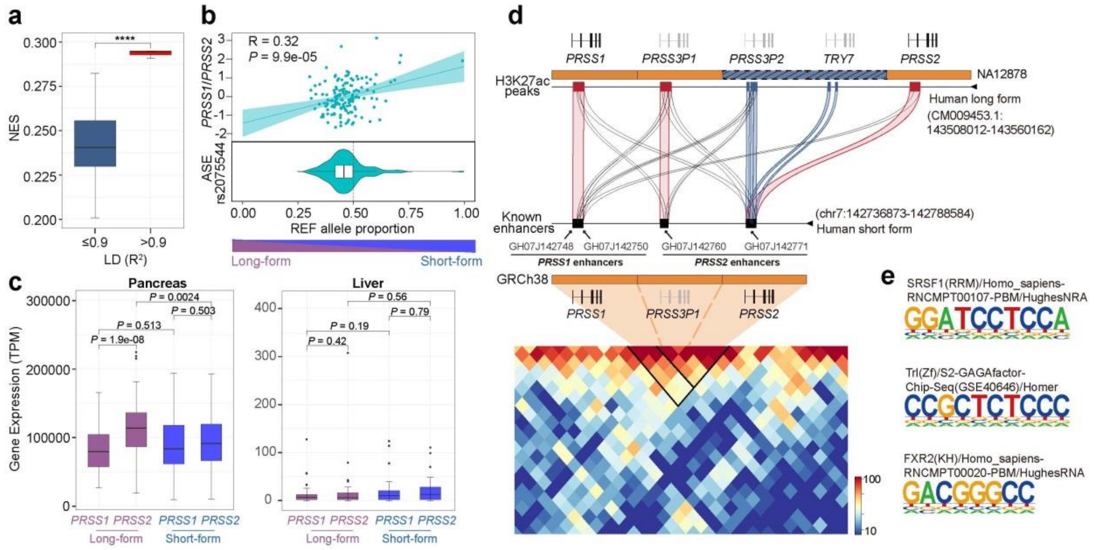


**Supplementary Fig. 19. Pseudogene enhancers account for increased *PRSS2* expression in long-form haplotype.**

**a.** Comparison of absolute normalized effect-size in *PRSS2* eQTL SNVs with different LDs of copy number structure (blue, $R^2 \leq 0.9$; red, $R^2 > 0.9$). **b.** Allele-specific expression (ASE) of copy number structure tagged SNV (rs2075544) in heterozygous 3-copy/5-copy samples. Lower panel: lower reference (REF, GRCh38 primary) allele proportion represents higher long-form (5-copy) haplotype expression, while higher reference (REF, GRCh38 primary) allele proportion represents higher short-form (5-copy) haplotype expression. Dash vertical line indicates the equal expression of long-form and short-form haplotype. Upper panel: The REF allele proportion is significantly correlated with *PRSS1/PRSS2* expression ratio. **c.** Gene expression of *PRSS1* and *PRSS2* in homologous short-form and long-form samples in pancreas (left) and liver (right). *PRSS1* and *PRSS2* showed significantly expression difference only in long-form haplotype in pancreas. **d.** Upper panel: Miropeats plot of sequence identity between the H3K27ac enhancers on the long-form and the known enhancers on the short-form haplotypes. The alignments with highest similarity are highlighted in color: red, the genes shared between long-form and short-form haplotypes; blue, the pseudogenes specific to the long-form haplotype. Lower panel: Hi-C contact map of homologous short-form haplotypes on *de novo* assembly CHM13 (3-copy structure) with resolution of 5-kb. **e.** Transcription binding site motif that are specific to *PRSS3P2*, *TRY7* and *PRSS2*, but absent in *PRSS1*.

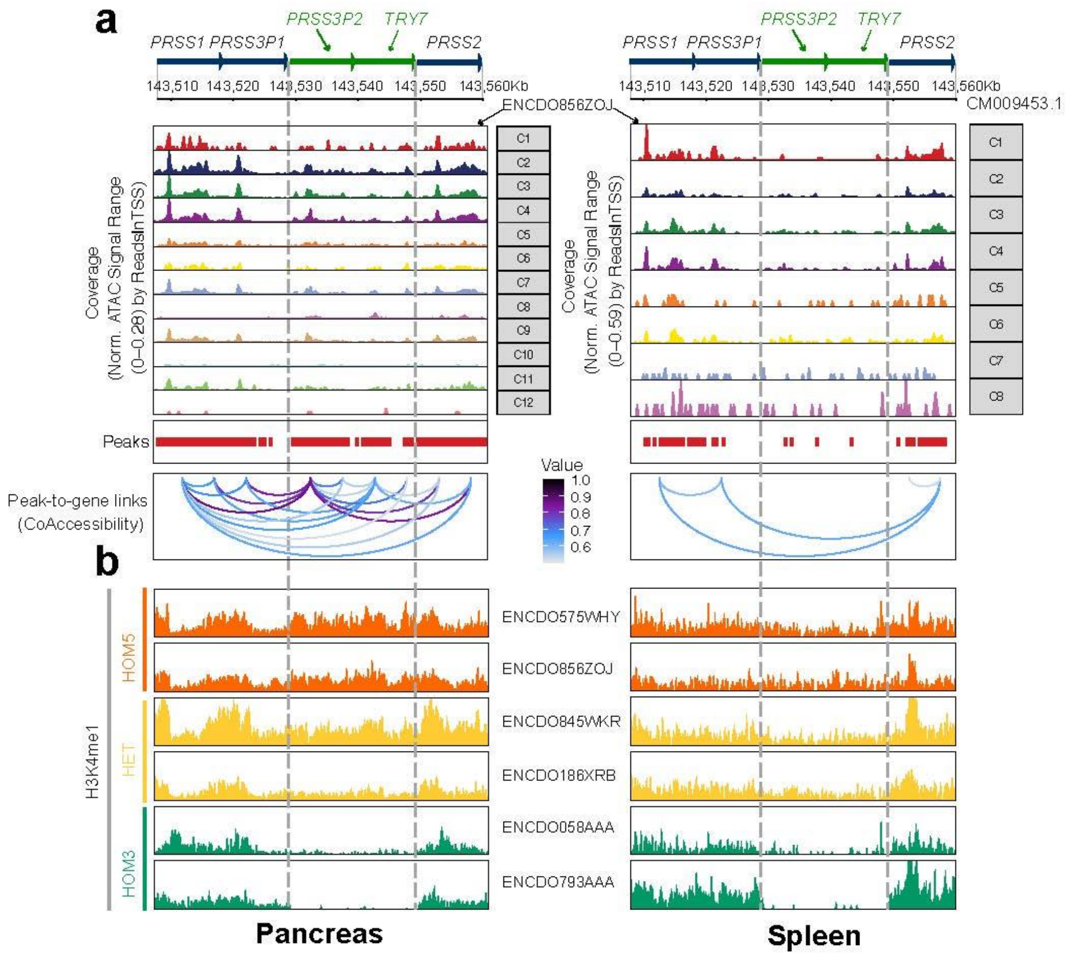


**Supplementary Fig. 20. Comparison of single-cell open chromatins states and histone modification signals between pancreas and spleen.**

**a.** Comparison of single-cell open chromatins states between pancreas and spleen for the same homologous long-form sample (ENCDO856ZOJ). Upper panel: the open chromatin states in different cell clusters. Lower panel: coaccesibility (loops) between the chromatin states and the genes. Noted that the peaks at polymorphic pseudogene copy region are weak and even disappear in spleen compared with those in pancreas. **b.** H3K4me1 histone modifications show difference signals at the polymorphic pseudogene copy region for the same samples in two tissues. Vertical dash lines indicate the polymorphic pseudogene copies that specific to long-form haplotype.

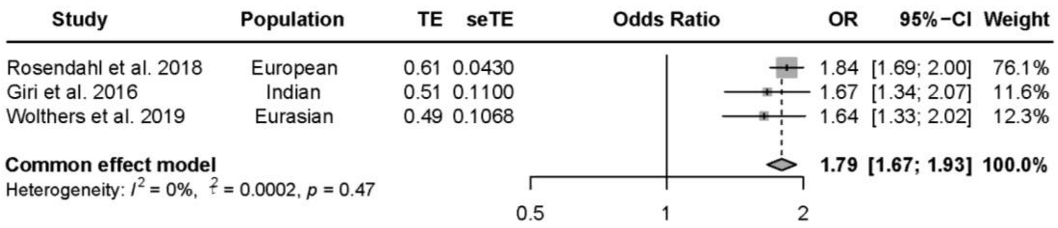


**Supplementary Fig. 21. Meta-analysis of association between copy number structure and pancreatitis risk.**

The variants included in this meta-analysis are in complete linkage disequilibrium with the copy number structure in non-African population. rs2855983 was used for ref. [6,7] and rs13228878 was used for ref. [8].

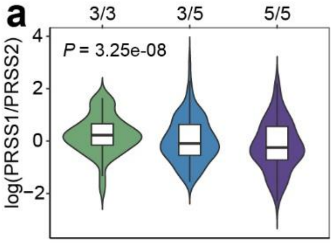


**b**

H0 : neither trait has a genetic association in the region
H1: only trait 1 has a genetic association in the region
H2: only trait 2 has a genetic association in the region
H3: both traits are associated, but with different causal variants
H4: both traits are associated and share a single causal variant

| Trait1 (GWAS) | Trait2 (Gene expression) | H0 | H1 | H2 | H3 | H4 |
|---|---|---|---|---|---|---|
| Pancreatitis (Rosendahl et al. 2018) | *PRSS1* | 1.995270e-33 | **0.987** | 7.329888e-36 | 0.004 | 0.009 |
| | *PRSS2* | 1.468507e-37 | 7.261560e-05 | 1.071935e-35 | 0.004 | **0.996** |
| | *PRSS1/PRSS2* | 5.662885e-38 | 2.800218e-05 | 7.151120e-36 | 0.003 | **0.997** |

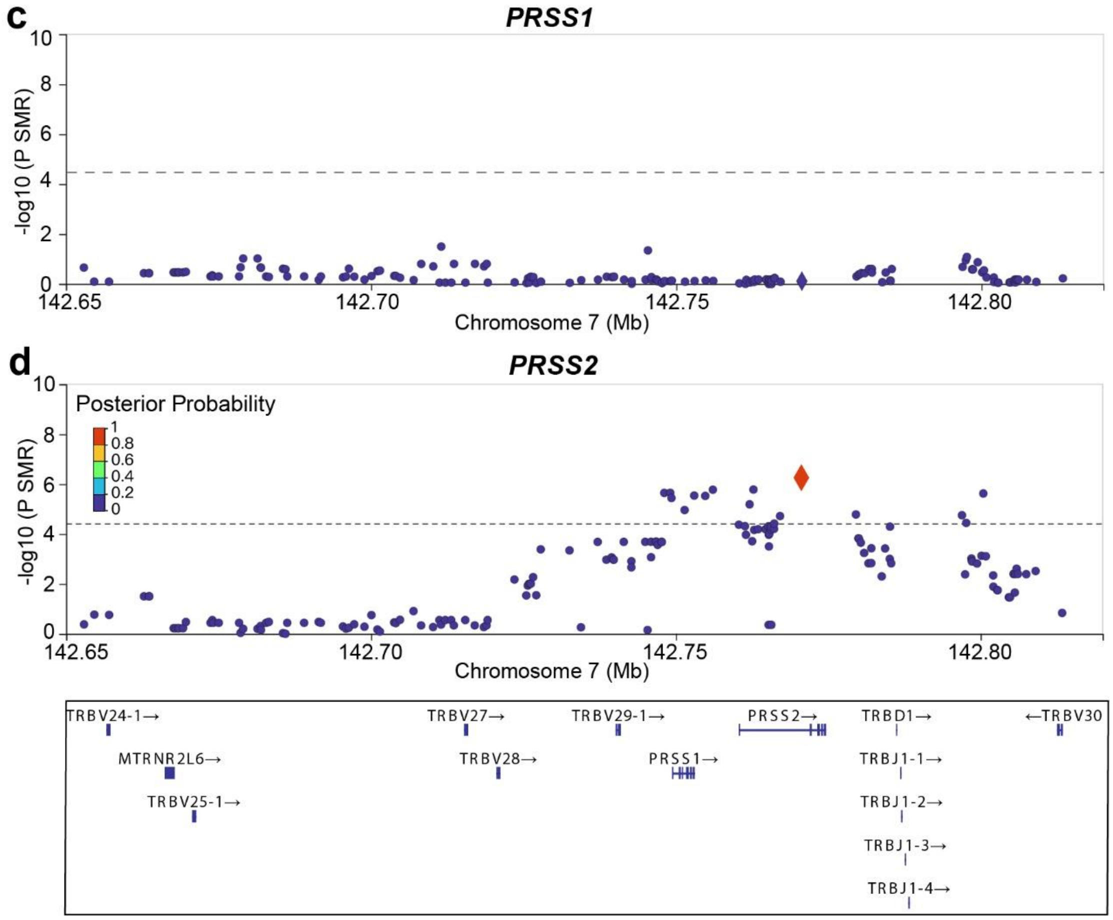


**Supplementary Fig. 22. Summary-data-based Mendelian Randomization and genetic colocalization analysis of gene expression and pancreatitis.**

**a.** Gene expression ratio (*PRSS1*/*PRSS2*) is significantly associated with copy number structure, in which the 5-copy form has lower ratio than the 3-copy form. **b.** Posterior probability (PP) under different hypothesis in the colocalization analysis of trait1 (pancreatitis from GWAS) and trait2 (gene expression). Only trait2 with *PRSS2* or *PRSS1*/*PRSS2* has high PP of H4, indicating two traits share the same causal factor. **c.** P-value of Summary-data-based Mendelian Randomization (SMR) for *PRSS1* expression and pancreatitis. The SMR analysis shows no significant signals suggest the effect size of genetic variants on pancreatitis is not mediated by *PRSS1* expression. **d.** SMR and colocalization analysis for *PRSS2* expression and pancreatitis. The significant SMR result suggests the effect size of genetic variants on pancreatitis is mediated by *PRSS2* expression, and the colocalization analysis shows the polymorphic pseudogenes are the causal factor for pancreatitis. In **c** and **d**, each circle dot represents a genetic variant, and the diamond denotes the variant of complete LD with copy number structure. Dash line indicate multiple-testing P-value cut-off. Color of the circle dot in **d** denotes the posterior probability of hypothesis H4.

### Supplementary Table Legends

**Supplementary Table 1. Coordinates of trypsinogen gene in primate genomes.**

**Supplementary Table 2. Human samples with inferred haplotypes included in this study.**

**Supplementary Table 3. Three-nucleotide motif used in characterizing haplotype groups.**

**Supplementary Table 4. Amino acid changes based on GRCh38 alternative contig.**
